# Enhancing Liver Fibrosis Measurement: Deep Learning and Uncertainty Analysis Across Multi-Centre Cohorts

**DOI:** 10.1101/2025.05.12.25326981

**Authors:** Marta Wojciechowska, Stefano Malacrino, Dylan Windell, Emma L. Culver, Jessica K. Dyson, the UKAIH consortium, Jens Rittscher

## Abstract

Digital pathology enables large multi-centre studies of histological specimens, but differences in staining protocols and slide quality can compromise the comparability of quantitative results. We analysed 686 PSR-stained liver biopsies from four independent cohorts spanning more than 20 clinical sites to assess how stain variability affects automated fibrosis quantification and model uncertainty. A U-Net ensemble was trained to segment collagen and to estimate pixel- and tile-level predictive uncertainty. Across markedly heterogeneous staining conditions, the ensemble achieved strong segmentation performance (Dice 0.83–0.90) and produced informative uncertainty maps that identified artefacts and out-of-distribution regions. Epistemic uncertainty values were typically below 0.002, providing a practical criterion for flagging unreliable predictions. Our results demonstrate that ensemble-based uncertainty estimation complements stain-standardisation efforts by quantifying prediction confidence directly from model outputs, improving the reliability and interpretability of collagen proportionate-area measurements across multi-centre datasets. This framework supports more trustworthy and reproducible digital-pathology workflows for fibrosis assessment and other histological applications.

**Graphical Abstract:** 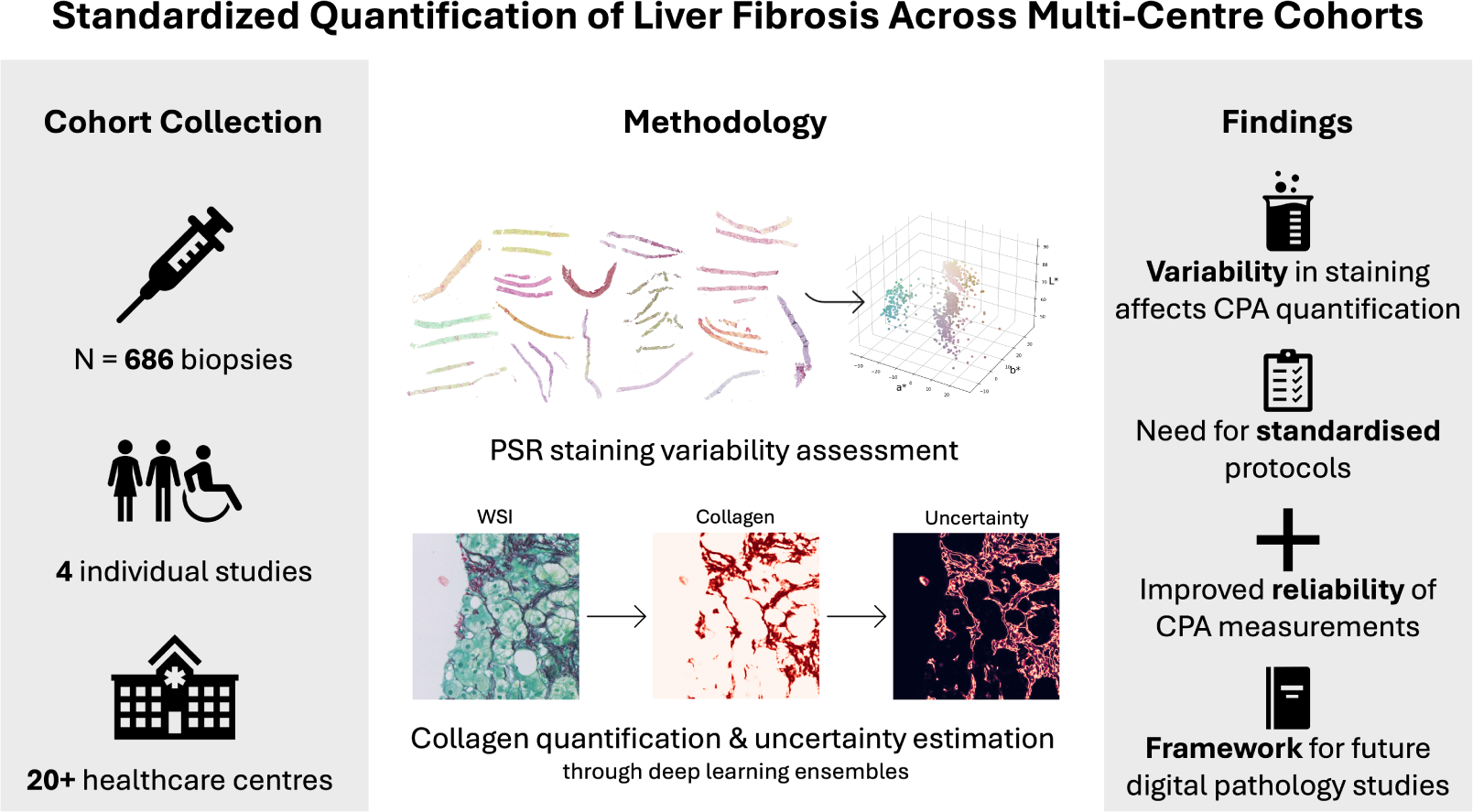

**Highlights:**

- A retrospective cohort of liver biopsies collected from over 20 healthcare centres has been assembled.
- The cohort is characterized on the basis of collagen staining used for liver fibrosis assessment.
- A computational pipeline for the quantification of collagen from liver histology slides has been developed and applied to the described cohorts.
- Uncertainty estimation is evaluated as a method to build trust in deep-learning based collagen predictions.

## 1. Introduction

Digital pathology enables scalable, multi-centre studies of histological biomarkers, yet systematic differences in slide preparation and image acquisition continue to hinder reproducible quantification. In this work we examine how staining heterogeneity affects automated collagen quantification and explore how ensemble-based predictive uncertainty can be used to interpret and manage unreliable measurements. Using PSR-stained liver biopsies collected from four independent cohorts across more than 20 clinical sites, we develop a deep-learning framework that combines stain characterisation with model-ensemble uncertainty estimation to improve the reliability and interpretability of collagen proportionate-area (CPA) measurements in liver fibrosis.

Fibrosis is increasingly recognised as a core functional component of the tissue microenvironment, and CPA is an established image-based metric derived from whole-slide images (WSIs) used in liver research [1]. CPA has been proposed as a trial endpoint in liver disease and is associated with long-term outcomes in metabolic-associated fatty liver disease and alcoholic liver disease [2–4]. However, CPA reflects only collagen burden rather than architectural patterns of fibrosis, and its clinical utility depends critically on consistent detection and quantification. Despite its promise, the application of CPA as an imaging biomarker faces substantial challenges. Published findings often lack consistency, largely due to variations in collagen detection and quantification methodologies [5], which undermine comparability across centres.

The literature on stain standardisation is extensive. Early approaches relied on stain deconvolution and stain-vector estimation [6–8], but assumptions such as white-light illumination limit their accuracy for whole-slide imaging [9]. Later methods introduced histogram-matching and colour-transfer techniques [10, 11] and, more recently, generative frameworks such as CycleGAN and related autoencoders [12–16]. These models can produce visually convincing stain transformations, but their biological fidelity and impact on quantitative analysis remain uncertain.

Automated quality-control tools such as HistoQC are highly effective for detecting slide-level artefacts [17], but they do not quantify inter-cohort staining variability or model confidence. To address these gaps, we combine colour-space analysis with ensemble-based uncertainty estimation, enabling both cohort-level assessment of staining heterogeneity and model-level assessment of prediction reliability.

This paper first presents cohort-level colour analyses (Section 2.4) that document staining heterogeneity, then describes the U-Net ensemble and uncertainty estimation approach (Section 3), and finally evaluates how uncertainty maps relate to segmentation performance and practical quality control (Section 4).

The following sections provide a detailed examination of the sources of measurement variability that motivate our approach.

### 1.1. Causes of uncertainty in CPA measurement

First, it must be acknowledged that any biopsy sample may not be representative of the examined organ as a whole. It is commonly estimated that a liver biopsy captures approximately 1:50 000 of the organ’s volume [18]. Given the known heterogeneity of fibrosis distribution in chronic liver disease, a single-slice CPA measurement may under- or overestimate total collagen content. This limitation affects all histology-derived metrics and predictions and has been addressed elsewhere [19]. While we focus here on quantifying uncertainty within digitised slide images, a full evaluation of sampling variability in CPA lies beyond the scope of this study. However, from the moment of biopsy acquisition, there are still a number of confounding factors that need to be considered to obtain a reliable measure of the collagen area in the analysed sample.

The process of CPA measurement can be broken down into three separate detection and quantification tasks: 1) collagen in-situ localisation (i.e. chemical staining), 2) the acquisition of the digital image and 3) the quantification of the stained area relative to the area of the tissue section. Each of these processes is subject to its own sources of error, which collectively contribute to the overall uncertainty of the measurement.

### 1.2. In-situ localisation of collagen

Liver fibrosis is a condition characterised by the pathological deposition of collagen fibres within the liver parenchyma, primarily in portal and perisinusoidal areas, and is therefore assessed using collagen-binding stains. Other connective tissue fibres, such as elastin fibres are also present in fibrotic structures. In comparison, their quantity is typically small[20]. Several stainings for identifying connective tissue in general and collagen specifically have been independently developed and traditionally used in pathology studies and in the clinic [5]. These include: PicroSirius Red (PSR), Masson’s Trichrome staining for connective tissue, van Gieson’s stain, orcein, and others. Each of these stainings is used in pathology to make the structure of connective fibres visible and can be used to visually assess the grade of fibrosis. However, the only histochemical stain which is known to be collagen-specific and so can be used to quantify collagen is Sirius Red.

PicroSirius Red (PSR) is a staining developed in the 1960’s with the purpose of selectively staining collagen in histology samples [21]. The primary agents in this staining are Sirius red F3B (CI 35780, Direct red 80) and picric acid (2,4,6-trinitrophenol, TNP). In the presence of picric acid, Sirius red F3B selectively binds with type I collagen (thicker collagen fibres), and type III collagen (thinner fibrillar collagen/reticulin fibres) [22]. Because of the selective staining for type I and III collagen fibres and its sensitivity to fine collagen strands, the PSR protocol is preferred for automatic quantification of collagen in histology slides. Another commonly used protocol for clinical assessment of liver fibrosis is Masson’s Trichrome (MT), in which collagen and other connective fibres are stained blue. In this paper, only Sirius red-stained slides were available for the development of digital pathology methods.

In spite of its wide-spread use as a collagen-specific stain, there isn’t a single standard protocol for PSR staining, which is used globally across institutions. The classical form of PSR staining includes only the mixture of Sirius Red and picric acid for collagen detection [21]. However, laboratories often add additional stains to the same tissue slice, to simultaneously examine other forms of tissue pathology. Commonly, two other stains may be added: a nuclear stain (a haematoxylin) and a cellular plasma stain (typically a green dye). This lack of PSR staining standardisation between healthcare institutions leads to very significant differences in the colour of the stained slides. This phenomenon can be well observed within the UK-AIH cohort where 20 NHS UK hospitals contributed liver FFPE slides prepared within their pathology laboratories (Fig. 1). The figure illustrates how deep the visual differences between PSR-stained slides can be when comparing samples collected at different sites.

**Figure 1:**
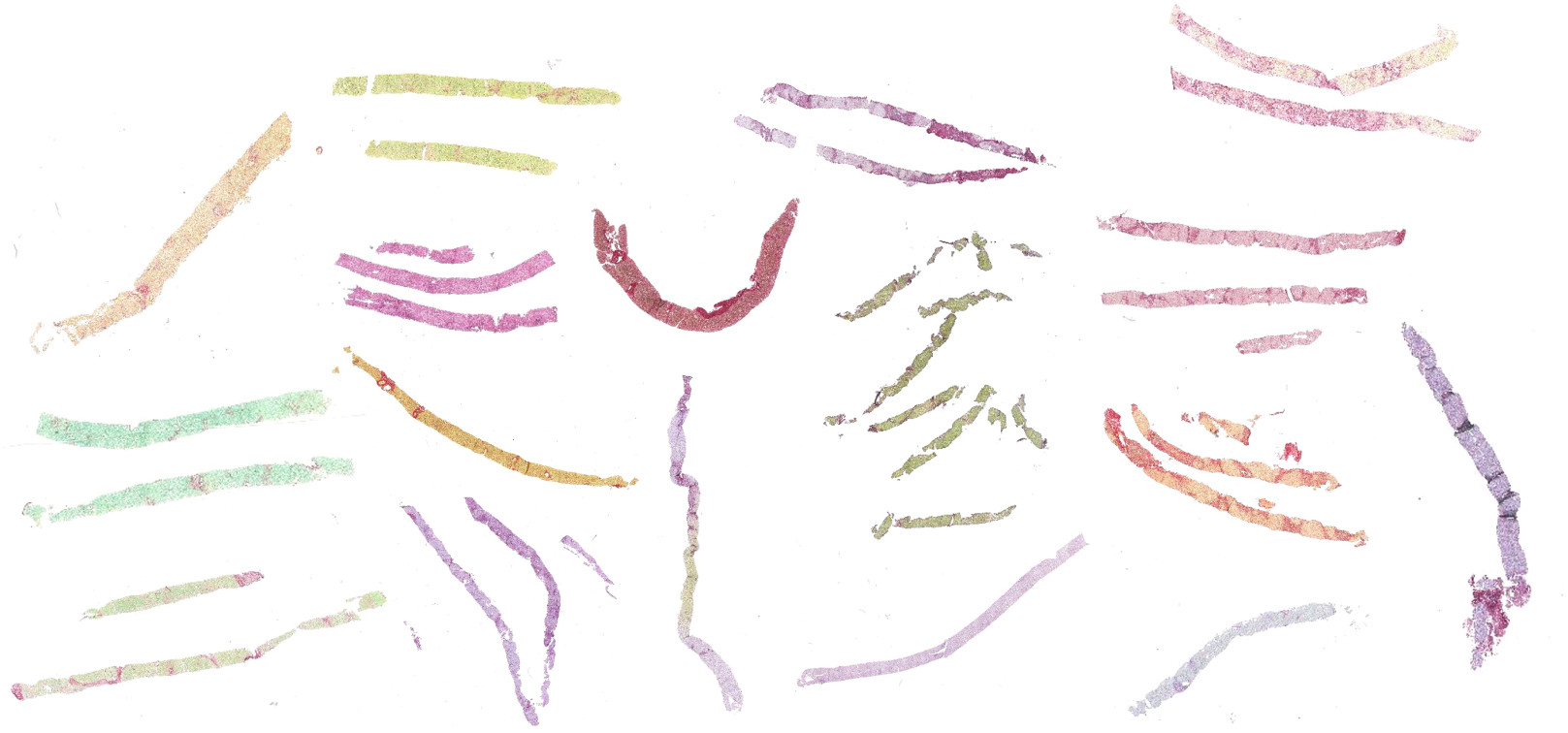
A visual representation of a selection of Sirius red-stained liver slides from the UK-AIH subcohort. This image illustrates how the use of different protocols for PSR collagen staining results in dramatic differences in slide appearance within the cohort collected retrospectively from 20 NHS hospitals. The images are not to scale. In each digital slide the background was removed and colours adjusted through histogram equalisation for visual clarity.

While PSR staining is known to create differential contrast between collagen strands and other tissue components, the exact sensitivity and specificity of PSR stain binding to collagen in a liver tissue section is not known [22]. Additionally, parameters such as the freshness of the specimen, stain concentration, stain exposure time, and ambient temperature can all affect the intensity of staining. It is not clear how the interactions of different chemicals and even the order in which different stains are applied affects the sensitivity and specificity of stain-collagen binding. Finally, histological stains are known to fade over the course of years and therefore retrospectively collected slides may not be comparable to freshly obtained ones in terms of colour intensity.

### 1.3. Digital sampling

The thickness of an individual collagen fibre is well below the *x,y* imaging resolution of a typical WSI scanner [23, 24]. Collagen fibres which can be seen in WSIs are aggregates of many collagen fibrils and are characterised by highly complex fractal-like shapes. Because of this fractal-like appearance of the analysed shape the measure of collagen area is intrinsically tied to the *x,y* resolution of the microscope objective lens and the size of the pixel of the imaging sensor [25]. The dependency of the quantified collagen area on the imaging magnification is significant. Which is why fractal metrics of dimensionality have been successfully applied to predict the overall stage of fibrosis [26].

Beyond the imaging resolution in the xy-plane, parameters such as slice thickness, lighting conditions and sensor exposure time directly affect the intensity of the colour of the digitised slide [27]. Clinical WSIs are typically acquired at either 20x or 40x magnification, which corresponds to *x,y* pixel sizes of approximately 0.5 x 0.5 µm, and 0.2 x 0.2 µm, respectively (see Tab. 2). In contrast, typical clinical biopsies are sliced at the thickness range between 4 and 10 µm. Hence a WSI pixel can contain a wide range of collagen thicknesses within the corresponding *z* volume of the tissue [28].

The final colour intensity of an imaged pixel is therefore a function of 1) the thickness of the tissue slice, 2) the thickness of the imaged collagen fibre in the tissue volume and 3) the relative intensity of collagen staining.

### 1.4. Segmentation errors

Finally, at any resolution the results of CPA quantification depend on the quality of the segmentation of collagen fibres. Fig. 2 illustrates the difficulty of annotating collagen on an example tile from a SR stained liver biopsy specimen. For now we focus on the data of the PREV study and only consider Sirius Red staining. The stain is present in the entire tissue section and can shown some spatial heterogeneity. Typically, collagen fibres have an intensely red colour. But it can be seen that other regions within the tissue parenchyma (cell cytoplasm and even more so cell nuclei) are also coloured pink. Unspecific staining is a factor that needs to be taken into account. Compression artifacts are another source of noise that needs to be considered. This illustrates why annotating collagen fibres in liver histology slides is burdened with high inter- and intra-observer variability [29].

**Figure 2:**
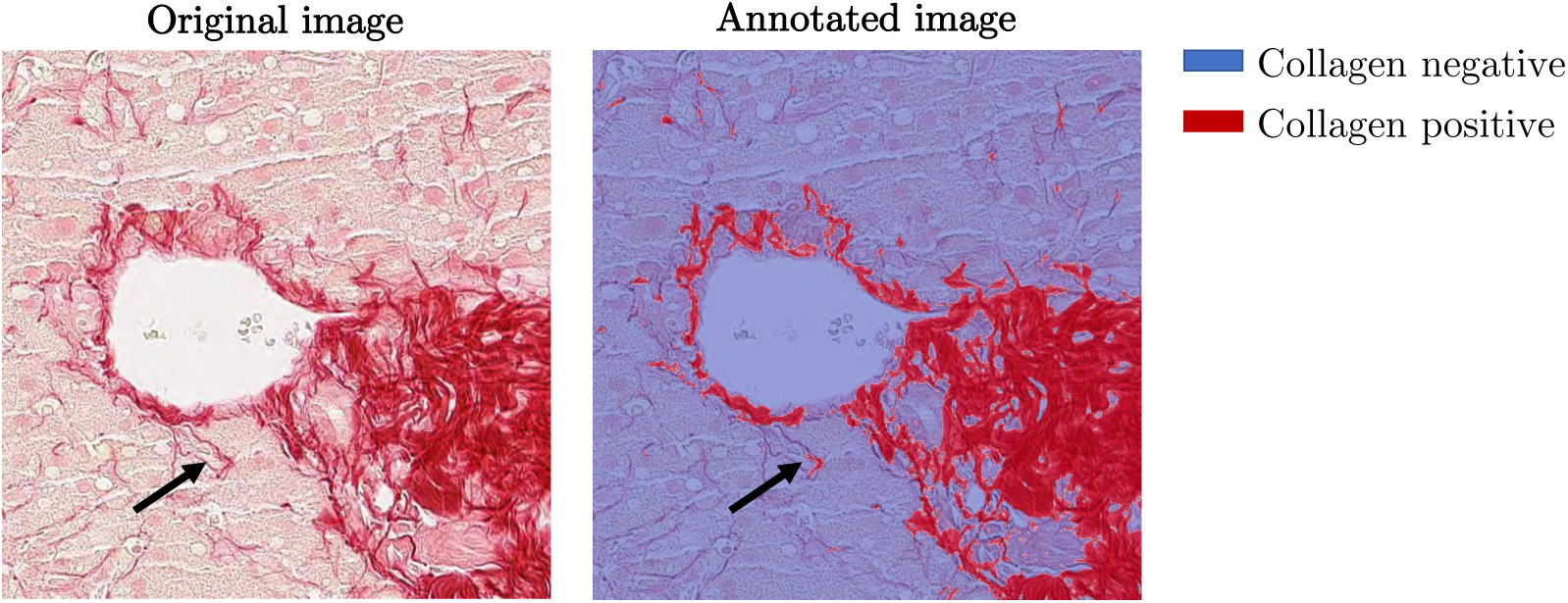
Limitations of creating human annotations of collagen fibres in microscopy images. The task is highly subjective and characterised by high interand intra-observer variability, particularly for thin collagen strands and in tissue slides with poor staining quality. While regions with thick, intensely stained collagen are captured, thinner fibres were missed by the annotator (MW).

Performing manual or even semi-automatic annotations of collagen area is a tedious task which can easily be automated by the employment of image analysis methods. Simple annotation tools often rely on the manual selection of a pixel intensity threshold by the annotating pathologist, such pipelines are known to be vulnerable to stain intensity gradients which are often found in histology slides. Deep learning segmentation models can be trained against pathologist segmentations to be robust to minor and even major differences in stain colour.

## 2. Materials and Methods

### 2.1. Cohorts and Data Acquisition

Four separate datasets of liver digital slides are used in this paper, henceforth referred to by the names of their respective original studies. Table 1 contains a brief summary of the cohort characteristics.

**Table 1:** An overview of the study cohorts.

| Cohort | N Slides | N Sites | Disease | Country |
| --- | --- | --- | --- | --- |
| CALM [30] | 153 | 2 | Mix | UK |
| HepaT1ca [31] | 44 | 1 | Liver Cancer, MAFLD | UK |
| PREV [32] | 279 | 1 | MAFLD | USA |
| UK-AIH [33] | 210 | 20 | AIH | UK |

In total, 686 digitised biopsies of mixed etiologies were gathered and analysed from a combined total of over 20 clinical sites. In each of the studies, liver biopsies or surgical resections were collected as part of standard care, with the exception of the PREV cohort which consists entirely of volunteers. Slides stained with H&E and PSR stainings were included in the analysis.

The samples within each of the three prospective cohorts in this paper: CALM, HepaT1ca and PREV have been processed according to internally agreed processing and staining protocols. The specimens from all of these studies have been stained with automatic stainers. In contrast to the above cohorts, the UK-AIH study is a retrospective study and slides dating back to 1998 from 20 hospital sites in the UK were selected. The FFPE slides in this cohort were processed according to internal standards within each of the participating hospitals at the time they were collected.

The CALM (ISCRTN39463479), HepaT1ca (NCT03213314) and PREV (NCT03142867) cohorts were made available by Perspectum Ltd., Oxford, UK. The UK-AIH cohort (IRAS ID: 144806) was made available by the UKAIH consortium funded by the NIHR Rare Diseases Translational Research Collaboration (RD-TRC) and coordinated by Newcastle University and Newcastle upon Tyne Hospitals NHS Foundation Trust. The UK-AIH study cohort was supported by LiverNorth, a UK-based patient support group and registered charity (No. 1087226). Further information about study design and data acquisition is available in the respective original publications [30–33].

### 2.2. Slide digitisation

The glass slides were digitised using whole slide imaging (WSI) scanners. Each cohort was scanned internally using parameters chosen by the individuals conducting the respective studies. Key parameters of the image acquisition are summarised in Table 2.

**Table 2:** Overview of digital slide parameters for each of the cohorts.

| Cohort | Scanner | Pixel Size: 20x<br>Magnification<br>[$\mu\text{m}$] | File<br>Format | Compression |
| --- | --- | --- | --- | --- |
| CALM | Hamamatsu<br>C12000-02 | 0.4533, 0.4533 | *.ndpi | JPEG *level*<br>*unknown* |
| HepaT1ca | Hamamatsu<br>C12000-02 | 0.4533, 0.4533 | *.ndpi | JPEG *level*<br>*unknown* |
| PREV | Aperio *model un-*<br>*known* | 0.4954, 0.4954 | *.svs | Aperio<br>JP2000 RGB |
| UK-AIH | Leica SCN400 | 0.5000, 0.5000 | *.scn | JPEG *level*<br>*unknown* |

Scanners from several WSI manufacturers were used for data acquisition. WSI systems from different vendors are equipped with different optical instruments and digital image sensors. These differences in hardware mean that the whole-slide images were acquired at slightly different resolutions, given the same nominal magnification (e.g. 20x). Similarly, scanning the same histology slide with two WSI scanners from different vendors produces digital images with slightly different colours [8]. This can be a result of both hardware differences and the colour calibration of a particular scanner. Each vendor also typically uses a proprietary image format.

All slides were processed using the vendor-neutral libraries: OpenSlide [34] and wsi-reader ^1^. Wsi-reader is a Python library developed by SM within the Rittscher group at the University of Oxford, designed to offer a unified interface for the most common WSI libraries, such as openslide, tifffile^2^ and Philips Pathology SDK.

### 2.3. Sirius red staining for collagen

To assess the stains used in each of the respective studies in the absence of staining protocols, stain deconvolution was performed on selected digitised slides from each cohort [6]. Stain deconvolution is a transformation of the image from the RGB colourspace into a colourspace in which each of the three image channels encodes the proportion of a colour of a single stain. This processing was performed using QuPath software [35]. While we cannot definitely identify particular stains or reconstruct staining protocols retrospectively using stain deconvolution, we are here using it to detect and document atypical colour profiles.

The stain deconvolution of the Sirius Red staining, shown in Figure 3, is used as an example to demonstrate the staining variability. Although a red collagen dye was used in each of the centres, it is obvious that different counter stains were used. This is in stark contrast to the results of stain deconvolution applied to the corresponding H&E slices from the same samples (Figure 4).

**Figure 3:**
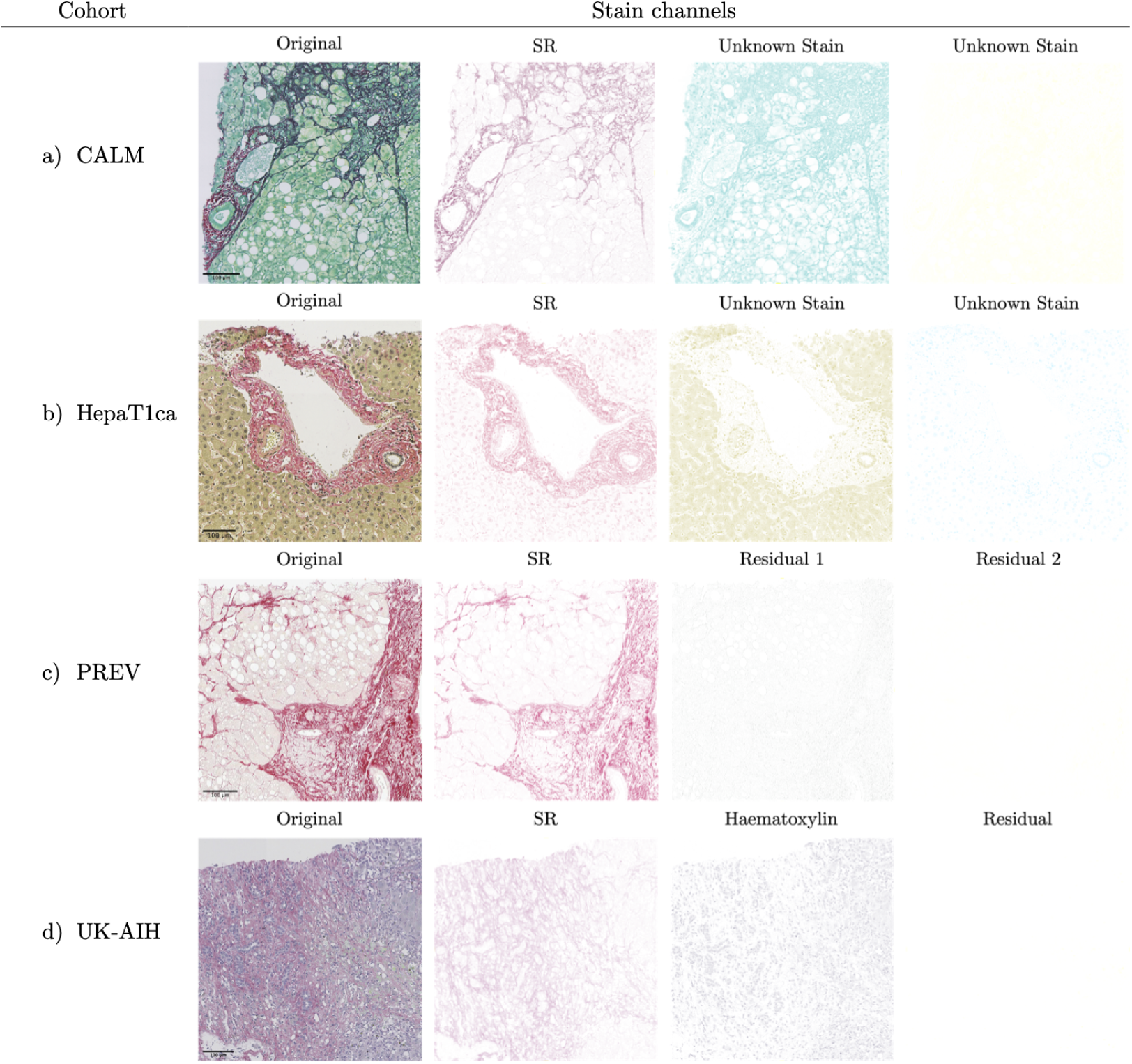
The results of stain deconvolution on example digital PSR slides from the analysed cohorts. a) CALM: the black colour of some collagen fibres is a result of non-specific binding of an unknown green stain to fibres stained with Sirius red. The slides contain a pale yellow residuum. b) HepaT1ca: Sirius red and two unknown stains, olive and blue. Upon close inspection one can see that cell nuclei are non-specifically stained red along with the collagen. c) PREV: only Sirius red stain was detected. d) UK-AIH: slide stained with Sirius red and a purple haematoxylin with an empty residual channel.

**Figure 4:**
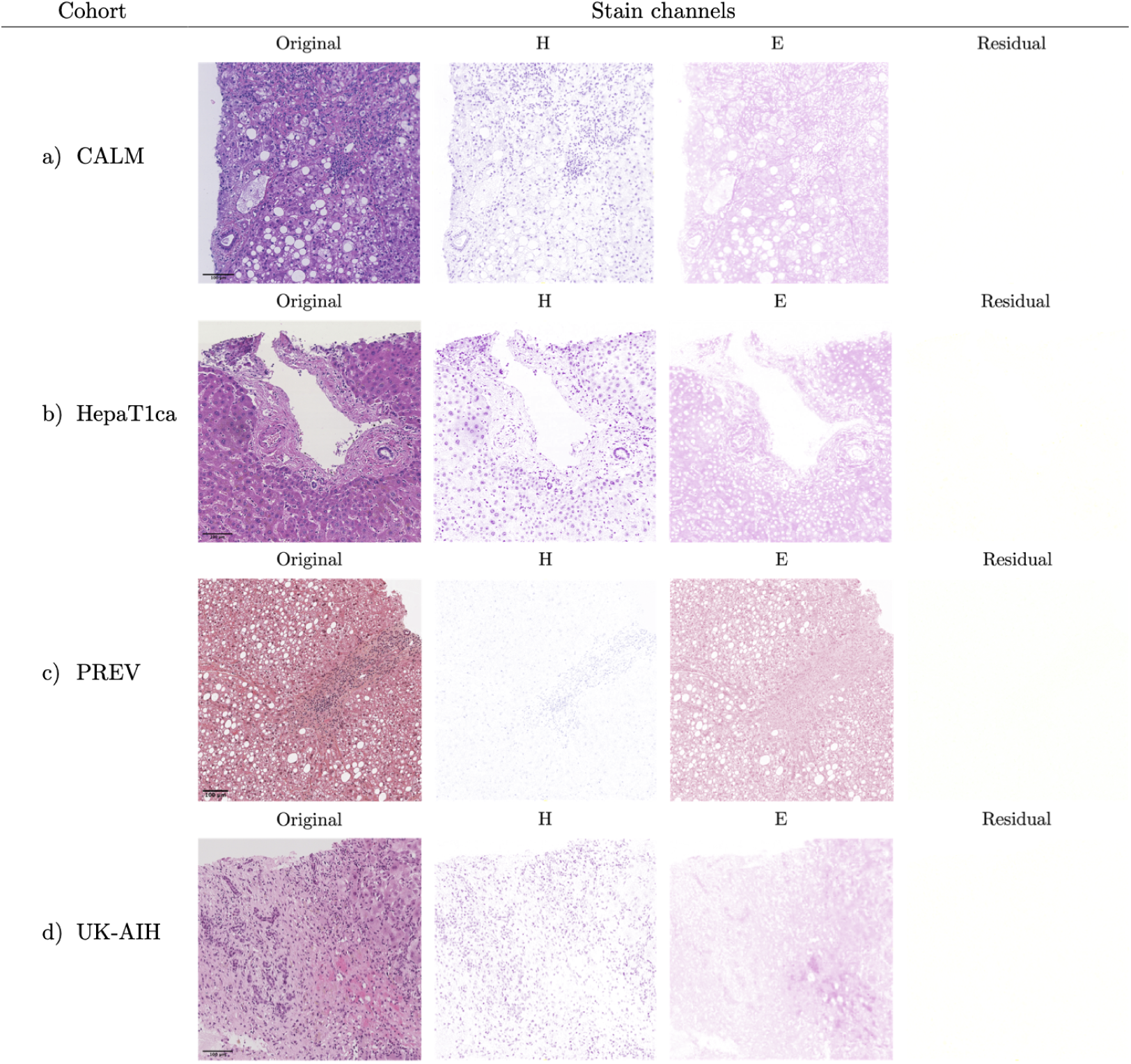
The results of stain deconvolution on example digital H&E slides from the four study cohorts. In each of the cohorts a haematoxylin and an eosin were used to produce H&E slides. The residual channel is empty or nearly empty in all of the slides. Slight differences in stain hue of both haematoxylin and eosin, as well as scan background colour can be seen between samples from different cohorts.

In the H&E-stained slides across all study cohorts is more standardised. In the PSR-labelled slides, the number of dyes used was ranging from one (just Sirius red) to three (i.e. Sirius red and two additional stains). In the case of the non-homogeneous UK-AIH cohort, different numbers and colours of stains were used for PSR staining.

### 2.4. Colour characterisation of digital slides

In order to visualise the spectrum of Sirius red-stained digitised FFPE section appearances, we here propose the concept of a primary colour of a digital slide (Fig. 5).

**Figure 5:**
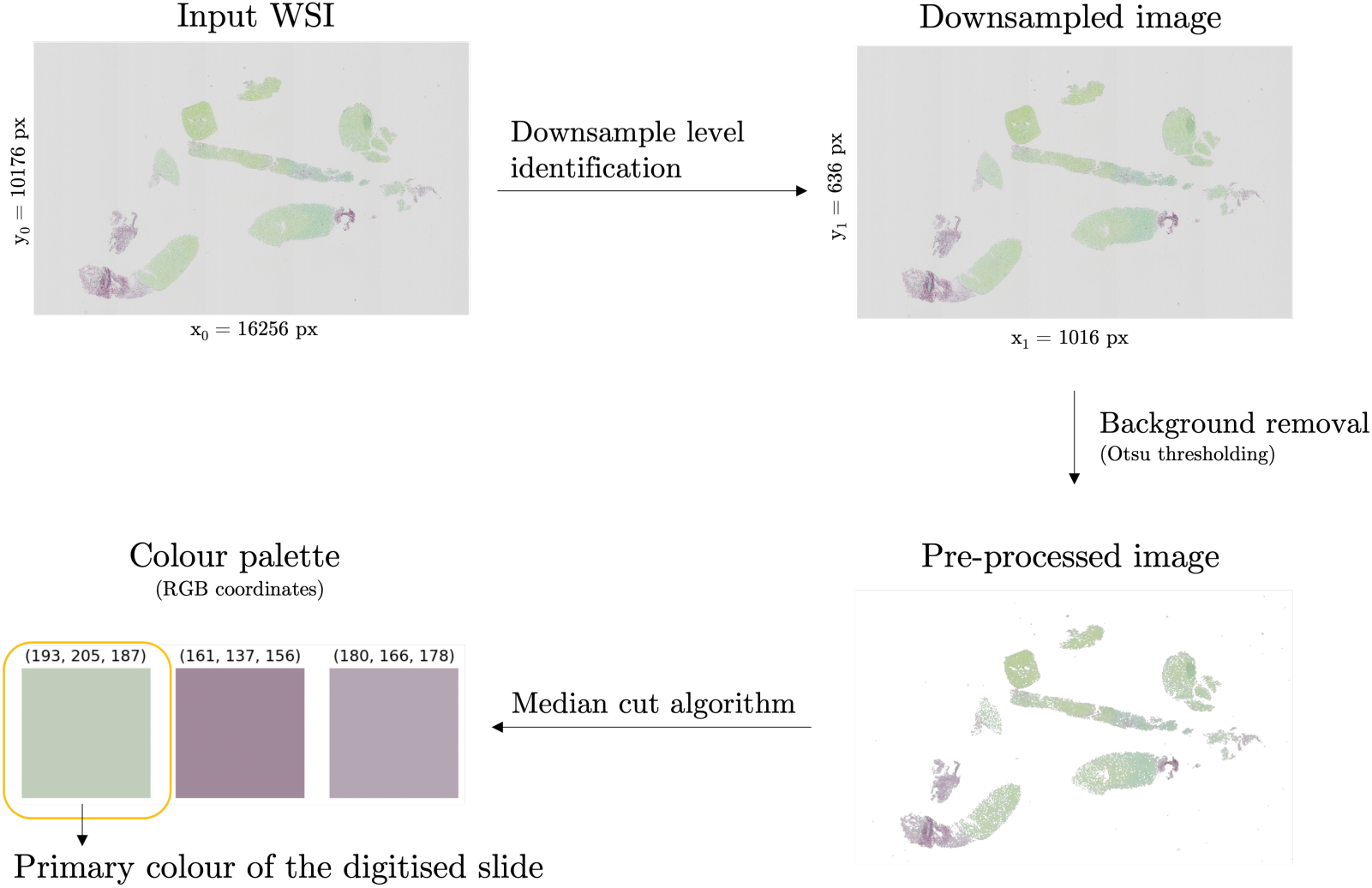
Identification of the primary colour of a digital slide. First, an appropriate downsample level is identified in the WSI pyramid. The slide image is then read from the downsampled level of the WSI file. Tissue/background intensity-based threshold is identified with the Otsu thresholding algorithm (applied to the greyscale-converted image). Background pixels are replaced with white values. Finally, the median cut algorithm is applied to thus pre-processed image to identify the primary RGB value in the digital slide.

One of the most commonly used approaches to quantisation of colours in digital images is through the employment of median cut algorithm [36]. Median cut algorithm is a method of sorting data of an arbitrary number of dimensions into series of sets by recursively cutting each set of data at the median point along the longest dimension. When applied to an image histogram, it outputs the most commonly occurring pixel values. Here, the WSI is first pre-processed by extraction of a downsampled version of the image and background removal. A pixel intensity threshold between tissue and background is established using the Otsu method applied to a grey-level version of the image [37]. All pixels of value above the established intensity threshold are replaced with white values. Median cut algorithm is applied to the pre-processed image to extract a three-colour colour palette, and the RGB values of the most commonly occurring colour encode the primary colour of the digital slide.

The primary colour has been computed for all of the SR and H&E-stained digitised slides available to this project. For purposes of comparison the RGB values of the primary slide colours were converted to the more perceptually uniform CIELAB colourspace [38]. The CIELAB colourspace expresses each colour as three values: L* for lightness, a* where negative values indicate green and positive values indicate red, and b* where negative values indicate blue and positive values indicate yellow. The results of primary colour estimation for the WSIs from all four cohorts are shown in Figures 6 and 7. These distributions quantitatively demonstrate the broad variability in Sirius Red staining across centres, complementing the qualitative examples in Figure 1.

**Figure 6:**
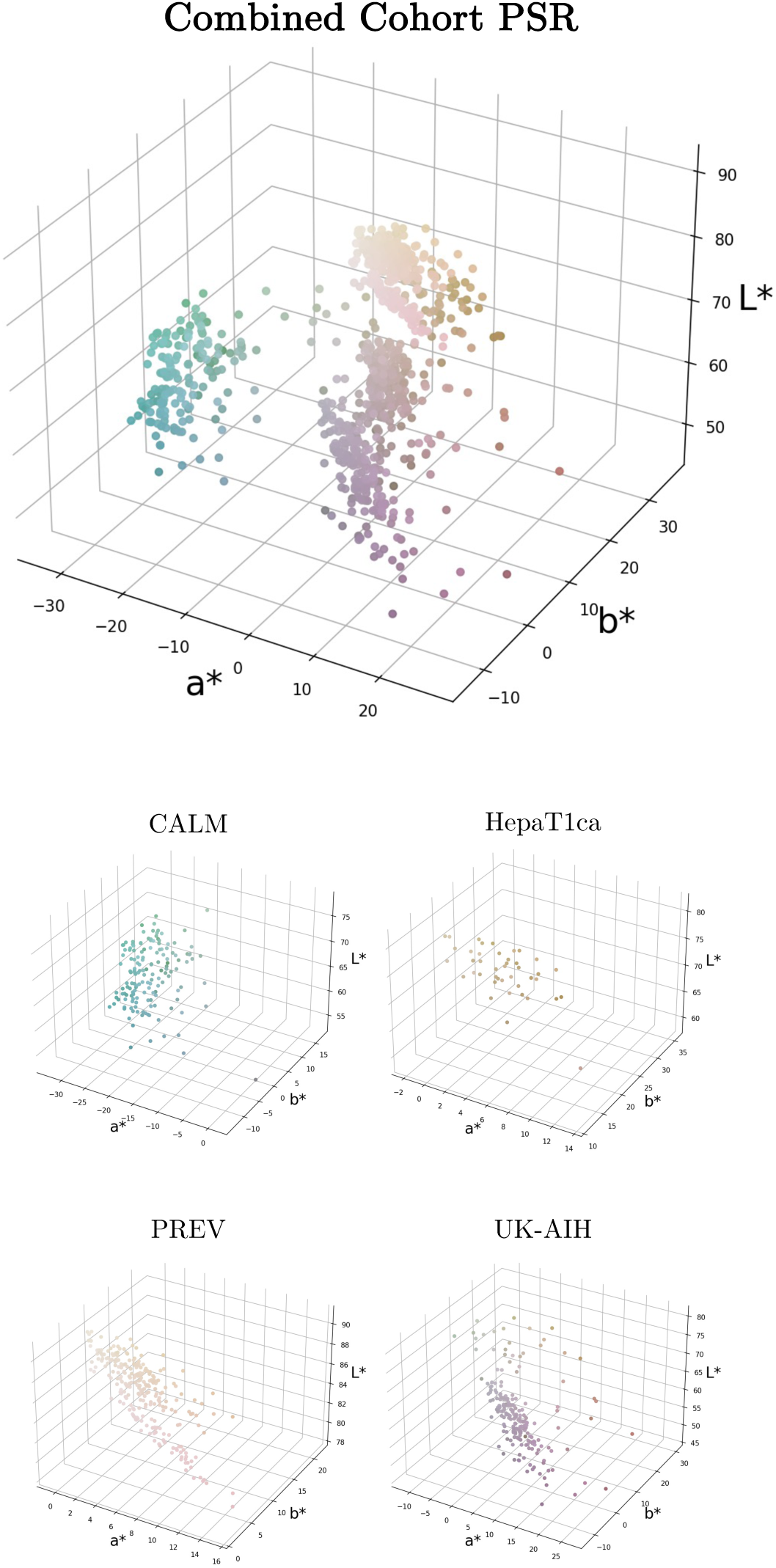
Primary colour distribution in digital slides of PSR-labelled sections. Samples from the individual cohorts form separate colour clusters.

**Figure 7:**
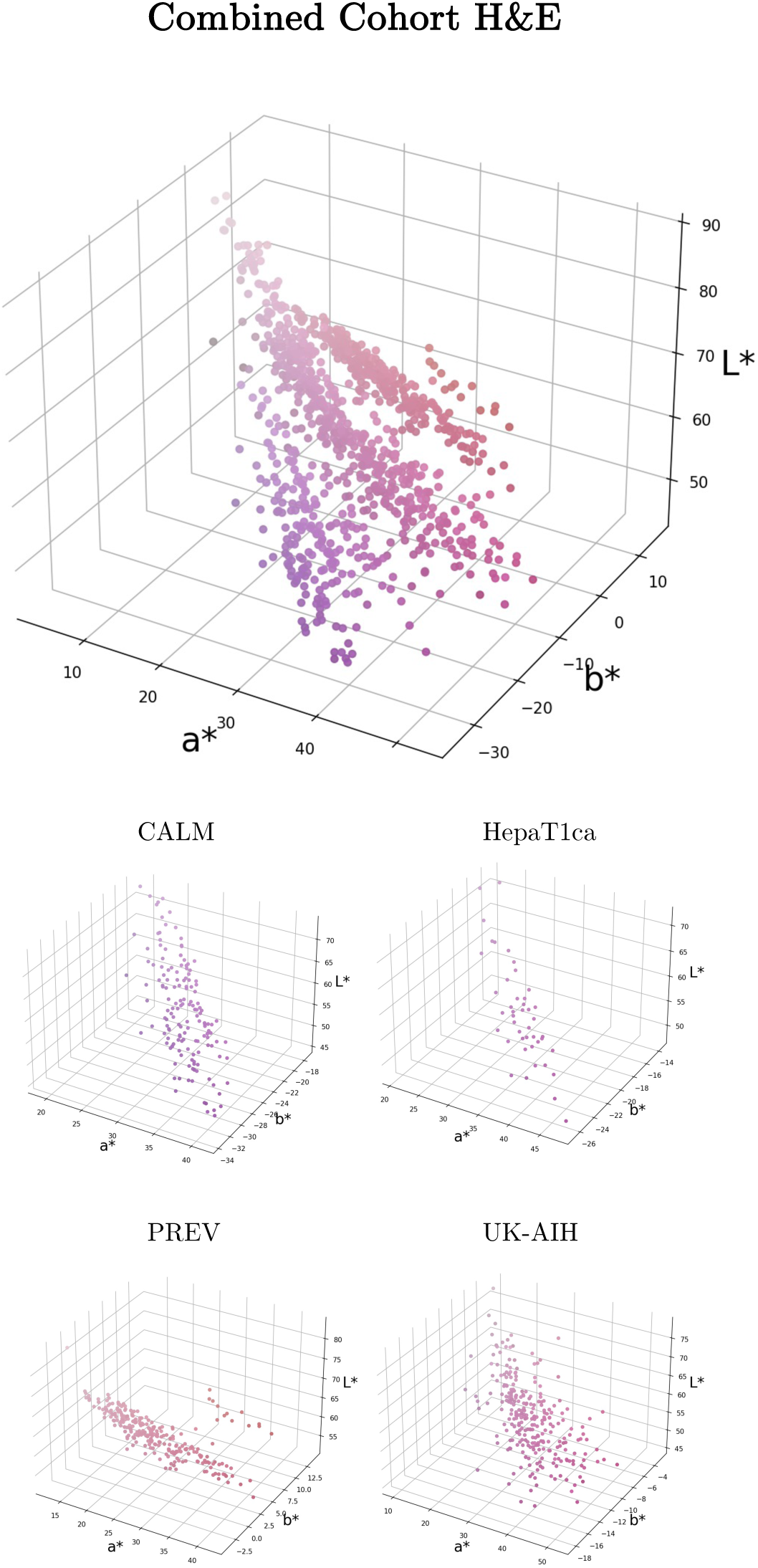
Primary colour distribution in digital slides of H&E-stained sections.

## 3. Computational Methods

### 3.1. Experimental Data

Table 3 summarises information about the training and validation data in this paper. Cases from the three training cohorts (CALM, PREV, UK-AIH) were manually selected using the obtained PSR stain distribution (Fig. 6) as a guide, to maximise the variation in colour in the experimental dataset. In total 38 whole slide images of liver biopsy sections were densely annotated by MW using QuPath software [35]. The pipeline for semi-automatic annotation of the slides with human in the loop is shown in Figure 8. Tiles containing less than 5% of annotated collagen area were discarded from the training and validation set to improve the stability of model training. All tiles from the HepaT1ca study were retained as an independent (unseen) validation set. A total of 2367 annotated 512×512 px tiles were used in the analysis. Of these 1970 were used for model training and 397 for model validation.

**Figure 8:**
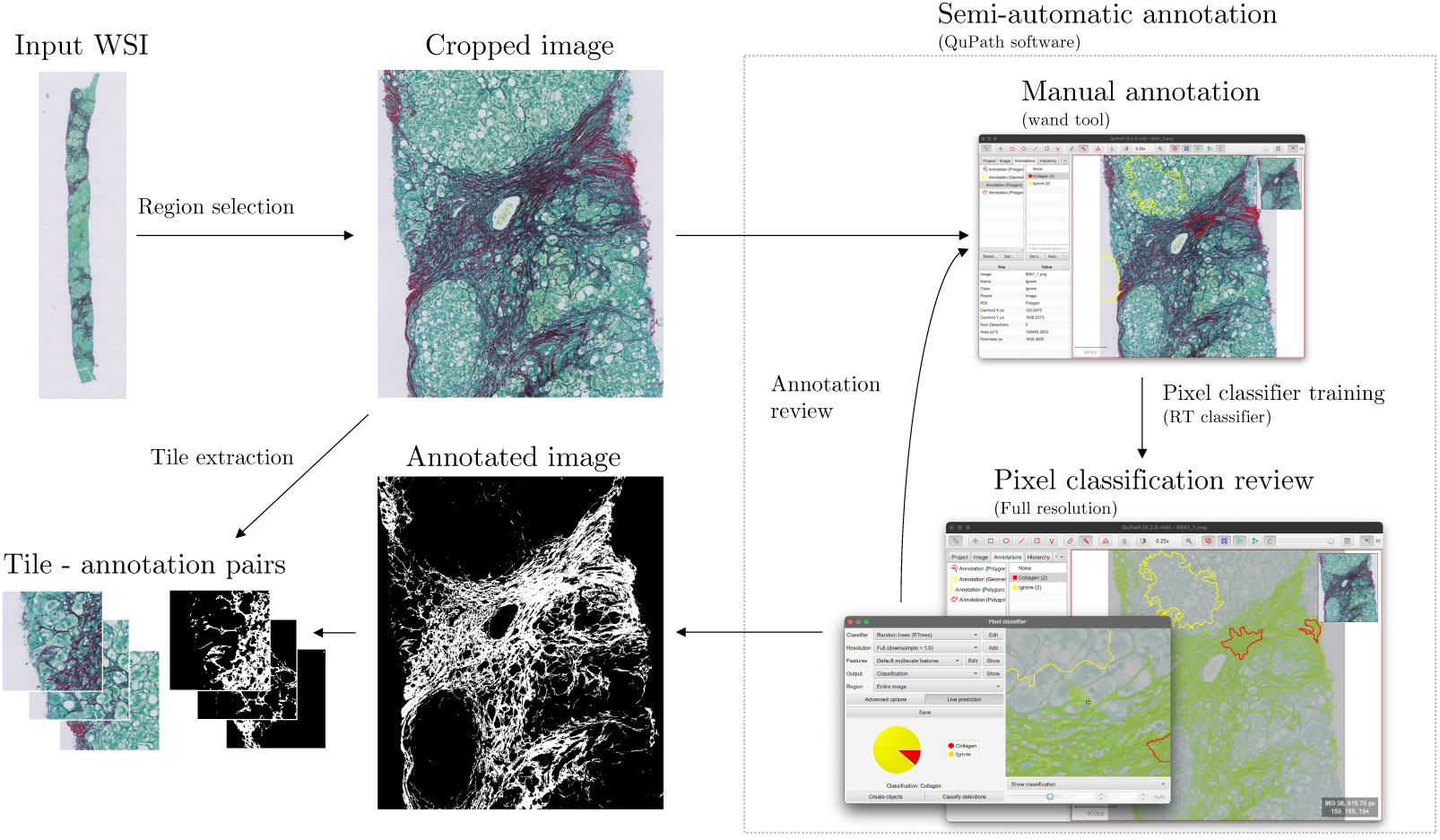
Collagen annotation with human in the loop. Firstly, a representative region is cropped from the full resolution level of the WSI file. The annotator trains a builtin random trees (RT) pixel classifier to categorise individual pixels within the image as either: “collagen” or “ignore”. The results of the RT classification are carefully examined visually. If needed, the human annotations are refined, and the process is repeated until the classifier produces a satisfactory pixel-level classification for the entire image. The annotation is then exported as a binary mask.

**Table 3:** A summary of training and validation data for each of the study cohorts. Images from CALM, PREV and UK-AIH cohorts were used for method development. HepaT1ca was retained as an independent validation dataset.

| Cohort | CALM | PREV | UK-AIH | HepaT1ca |
| --- | --- | --- | --- | --- |
| N Annotated Cases | 12 | 13 | 13 | 5 |
| N Train Tiles | 442 | 1172 | 356 | - |
| N Validation Tiles | 53 | 141 | 52 | 151 |
| Mean Collagen % | 21% | 15% | 24% | - |

### 3.2. Segmentation Models

The U-Net is a popular deep learning model used for medical image segmentation [39]. We trained three variants of the U-Net model for the collagen segmentation task using the implementation available at ^3^. The first two models (here called U-Net Tiny and U-Net Mini) are each down-sized versions of the original U-Net. An Attention U-Net [40] was trained for the same task. The parameters of the individual networks are summarised in Table 4.

**Table 4:**
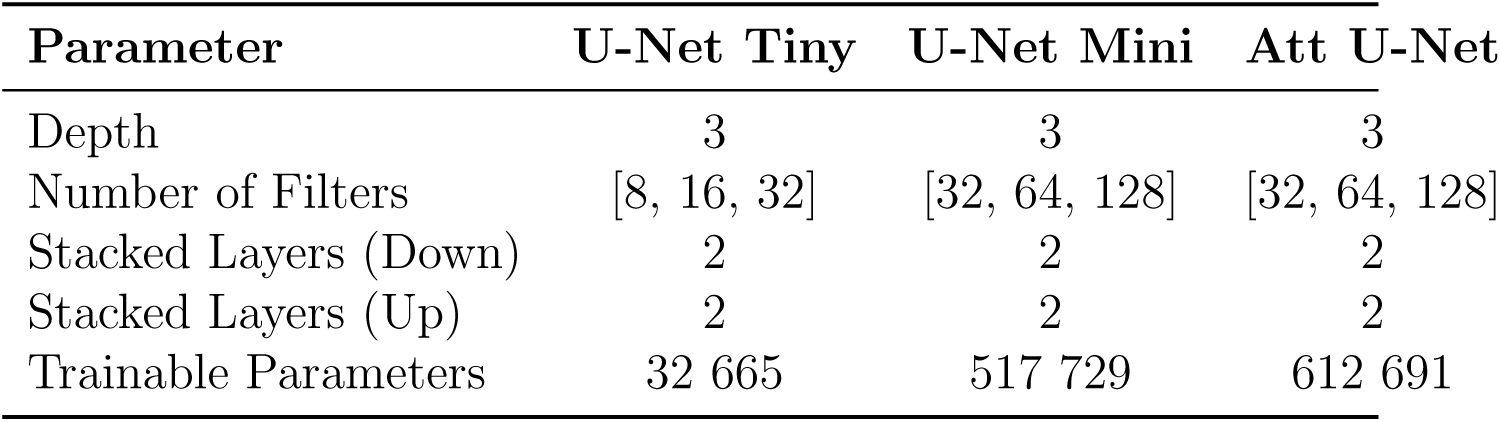
A summary of the parameters of the trained segmentation models.

| Parameter | U-Net Tiny | U-Net Mini | Att U-Net |
| --- | --- | --- | --- |
| Depth | 3 | 3 | 3 |
| Number of Filters | [8, 16, 32] | [32, 64, 128] | [32, 64, 128] |
| Stacked Layers (Down) | 2 | 2 | 2 |
| Stacked Layers (Up) | 2 | 2 | 2 |
| Trainable Parameters | 32 665 | 517 729 | 612 691 |

Each of the trained segmentation networks has an input which matches the size of 512×512 px RGB image tiles. The ReLU activation function was used in each segmentation model. U-Net Tiny and U-Net Mini used max pooling as the pooling strategy and a sigmoid activation function at the network output. Each of the models used bilinear unpooling. The models were trained using binary crossentropy loss and Adam optimiser. Each of the networks was trained from scratch with a randomised initialisation seed and a batch size of 8. A Quadro RTX 6000 GPU core was used for all of the experiments conducted.

### 3.3. Uncertainty Estimation

In order to estimate the real uncertainty of PSR measurement, one would need to perform an uncertainty estimation for each of the steps in sample processing and image acquisition separately. Unfortunately, as only digitised slides were made available to this project, such an analysis goes beyond the scope of this paper. Here we propose a way of estimating the uncertainty inherent to the digital image itself, which can be inferred by the image segmentation deep network.

Uncertainty estimation in the context of deep learning can improve the quality of predictions [41, 42]. Bayesian inference is a statistical framework which can be used to quantify the uncertainty present in deep-learning models. The concept is based on Bayes’ theorem, which links the posterior distribution of our observations to the prior probability of the hypothesis and the likelihood of the data given the hypothesis:

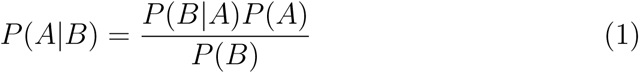

where: P(A—B) is the posterior probability of hypothesis A given the observed data B, P(B—A) is the likelihood of the observed data B given hypothesis A, P(A) is the prior probability of hypothesis A and P(B) is the marginal likelihood of the observed data B.

In the Bayesian framework can be used to quantify uncertainty. However, exact Bayesian inference with deep neural networks is computationally intractable. It can be approximated by Bayesian Neural Networks (BNNs), which are neural networks with probabilistic weights and biases. In the BNN framework, the weights and biases of a network are treated as random variables with a prior distribution over their values. Bayesian inference is then used in the process of training the network to update the prior distribution with new data. The posterior distribution over weights and biases obtained during network training can be used to compute the predictive distribution of the model. This predictive distribution of the model is a representation of the range of possible model outputs given an input. The uncertainty of the model can be inferred from the model’s predictive distribution and can be a measure of the model’s confidence in the predictions. An alternative approach to Bayesian neural networks is Bayesian model averaging, in which predictions of multiple models are averaged. In practice, several methods of applying Bayesian inference to uncertainty estimation in deep learning have been proposed. Those include Markov chain Monte Carlo [43], variational inference [44], Monte Carlo dropout (MC dropout) [41] and deep ensembles [45].

Predictive uncertainty can be decomposed into two components: aleatoric, which captures inherent randomness in the data (such as noise in the labels), and epistemic, which represents the uncertainty of the model (e.g. when making predictions on out-of-distribution samples) [46]. Kendall et al. [42] proposed an uncertainty decomposition method using a neural network that outputs two values representing the mean and variance of the prediction and using Monte Carlo dropout to approximate Bayesian inference. A similar approach was used by Kwon et al. [47] who proposed a new method for estimating aleatoric and epistemic uncertainties using Monte Carlo dropout. Fort et al. [48] recently demonstrated that deep learning ensembles with random initialisations allow for a more robust approach to uncertainty estimation. This is attributed to the fact that deep ensembles explore entirely different modes, while methods which are based on exploring the subspace of a single model (such as MC dropout) converge to a single mode.

Following [45], we used deep ensembles as an alternative to Bayesian inference. In our experiments, all models were trained for the same duration of 30 epochs. We trained separate ensembles of M = 10 models for each of the studied cohorts and an additional one for the combined cohort (CALM + PREV + UK-AIH). Overall 12 ensembles of M = 10 models each were trained. Slides from the HepaT1ca cohort were retained as a separate validation set.

Given an ensemble Ω of *M* trained models *Ω = {ω_m_}^M^_ω=1_*, we computed the pixel-wise collagen likelihood as the average *p*^ of the ensemble predictions *p_m_*for an input pixel *i*.

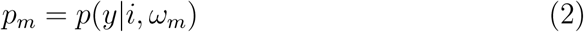

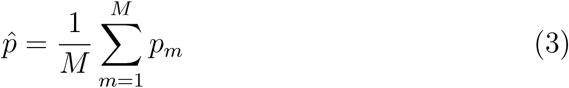

We then used the method proposed by [47] to estimate the pixel-wise predictive uncertainty for an input pixel *i* and the output prediction *y* as the variance of *y* given *i* over the predictive distribution *p*(*y|i,* Ω):

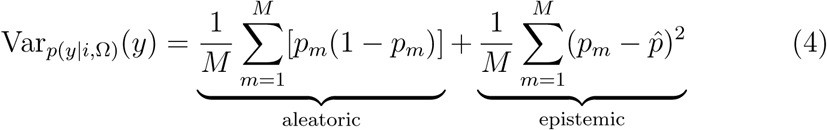

Two approaches for computing collagen percentage area in a tile were employed; the first made use of the mean ensemble prediction *P* and the other of the voted ensemble segmentation *B*. The mean collagen prediction *P* for image *I* with dimensions *W, H* is given by the equation:

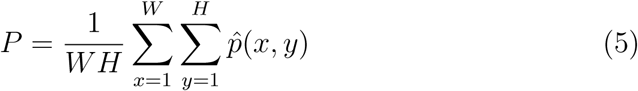

Collagen segmentation value ^^^*b* was obtained for each pixel via thresholding of the corresponding ensemble prediction at collagen likelihood equal or higher than 0.5. Let *p*^(*x, y*) be the ensemble prediction value for pixel *i* at location (*x, y*) in the image *I*, and let ^^^*b*(*x, y*) be the corresponding binary value after thresholding. Hence ^^^*b*(*x, y*) can be defined as:

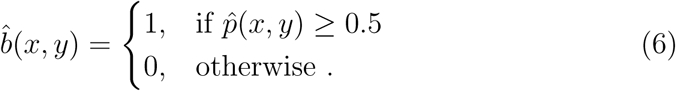

The segmented collagen area proportion B is given by:

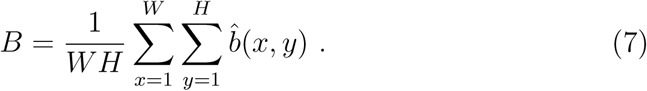

The uncertainty in the mean tile collagen prediction is reported as the interquartile range (IQR) of the ensemble collagen predictions *P*. Similarly, the uncertainty of the segmented collagen area in a tile is reported as the IQR of the collagen segmentation maps produced by thresholding of the predictions by each of the models in the ensemble.

## 4. Results

The validation metrics for the trained segmentation models are summarised in Table 5. We provide the scores separately for ensembles trained and validated on the individual study cohorts and for the ensemble models trained on the pooled cohort (CALM + PREV + UK-AIH).

**Table 5:** Segmentation network Dice scores on validation tiles. (M = 10 ensemble average ± standard deviation).

| Training Set | CALM | PREV | UK-AIH | HepaT1ca |
| --- | --- | --- | --- | --- |
| **Individual Cohorts** |  |  |  |  |
| U-Net Tiny | $0.847 \pm 0.022$ | $0.877 \pm 0.012$ | $0.793 \pm 0.036$ | - |
| U-Net Mini | **$0.905 \pm 0.026$** | **$0.882 \pm 0.005$** | **$0.852 \pm 0.019$** | - |
| Att U-Net | $0.820 \pm 0.022$ | $0.825 \pm 0.059$ | $0.759 \pm 0.070$ | - |
| **Pooled Cohort** |  |  |  |  |
| U-Net Tiny | $0.823 \pm 0.021$ | $0.861 \pm 0.013$ | $0.767 \pm 0.037$ | **$0.594 \pm 0.075$** |
| U-Net Mini | **$0.834 \pm 0.029$** | **$0.864 \pm 0.014$** | **$0.795 \pm 0.035$** | $0.555 \pm 0.104$ |
| Att U-Net | $0.785 \pm 0.036$ | $0.834 \pm 0.017$ | $0.623 \pm 0.067$ | $0.539 \pm 0.123$ |

The U-Net Mini has achieved the highest validation metric scores in all of the assigned tasks. It can be seen that for all model architectures there is a drop in Dice scores between the ensembles trained on individual cohorts and those trained on the pooled cohort. The higher Dice scores for individual cohorts can be explained by the models over-fitting to the data within each of the cohorts. It can be seen that while very high Dice scores are achieved for the CALM and PREV cohorts, performance of each of the network architectures is slightly worse for the UK-AIH cohort. This can be explained by the fact that the UK-AIH samples were collected retrospectively from a large number of hospitals and are characterised by frequent staining artefacts and large intra-cohort differences in slide colour as shown in Figure 1 and quantitatively in Figures 6 and 7. As expected, the performance of the segmentation models is significantly lower for the unseen cohort (HepaT1ca). This drop in Dice scores is likely driven by cohort-specific differences in staining, colour profiles, and imaging protocols not represented in the training data. While our ensemble models were trained to capture variability across multiple centres, HepaT1ca remains an external dataset with distinct characteristics. This performance drop highlights the limits of generalisability even in diverse training settings and motivates future exploration of robustness-enhancing techniques.

Figure 9 shows model predictions for an example tile from the CALM cohort. It can be seen that the human annotation used as ground truth for model validation contains noisy false positive pixels around the annotated fibres. These noisy labels can be attributed to falsely positive labelling by the annotator (MW) of the JPEG compression-induced noise in the original WSI. All prediction maps generated by the tested U-Net models provide a realistic representation of the collagen pattern in the original image. It can be seen that neither of the trained segmentation models has replicated the image compression noise in the final prediction, which is a desirable outcome. It is difficult to notice any visual differences in the quality of the prediction maps output by each of the models. This shows that each of the tested networks is appropriate for the task of collagen segmentation.

**Figure 9:**
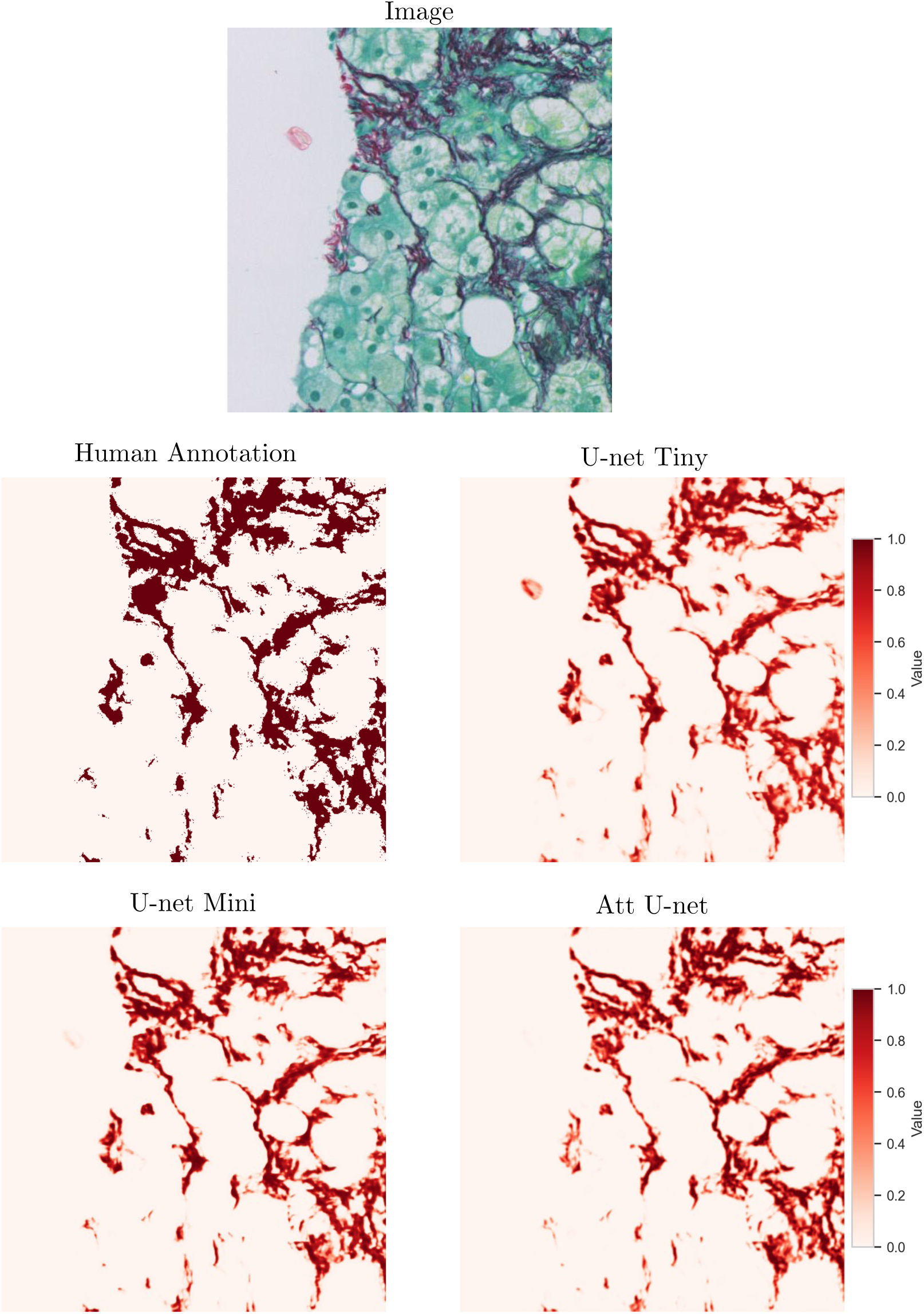
Collagen prediction maps output by the tested segmentation U-Net models. The human annotation used as ground truth for model validation contains noisy false positive pixels around the annotated fibres. Each prediction map shows an output prediction from a single model trained on the pooled cohort. It is difficult to see any differences in the quality of the collagen maps predicted by different model types.

### 4.1. Uncertainty computation

Our ensemble-based uncertainty estimates follow the framework introduced by Lakshminarayanan et al. [45], which demonstrated that deep ensembles provide a practical and well-calibrated alternative to Bayesian inference for predictive uncertainty estimation. In this context, the ensemble variance visualised in Figures 10–11 offers a qualitative view of calibration behaviour analogous to reliability plots used in classification tasks.

**Figure 10:**
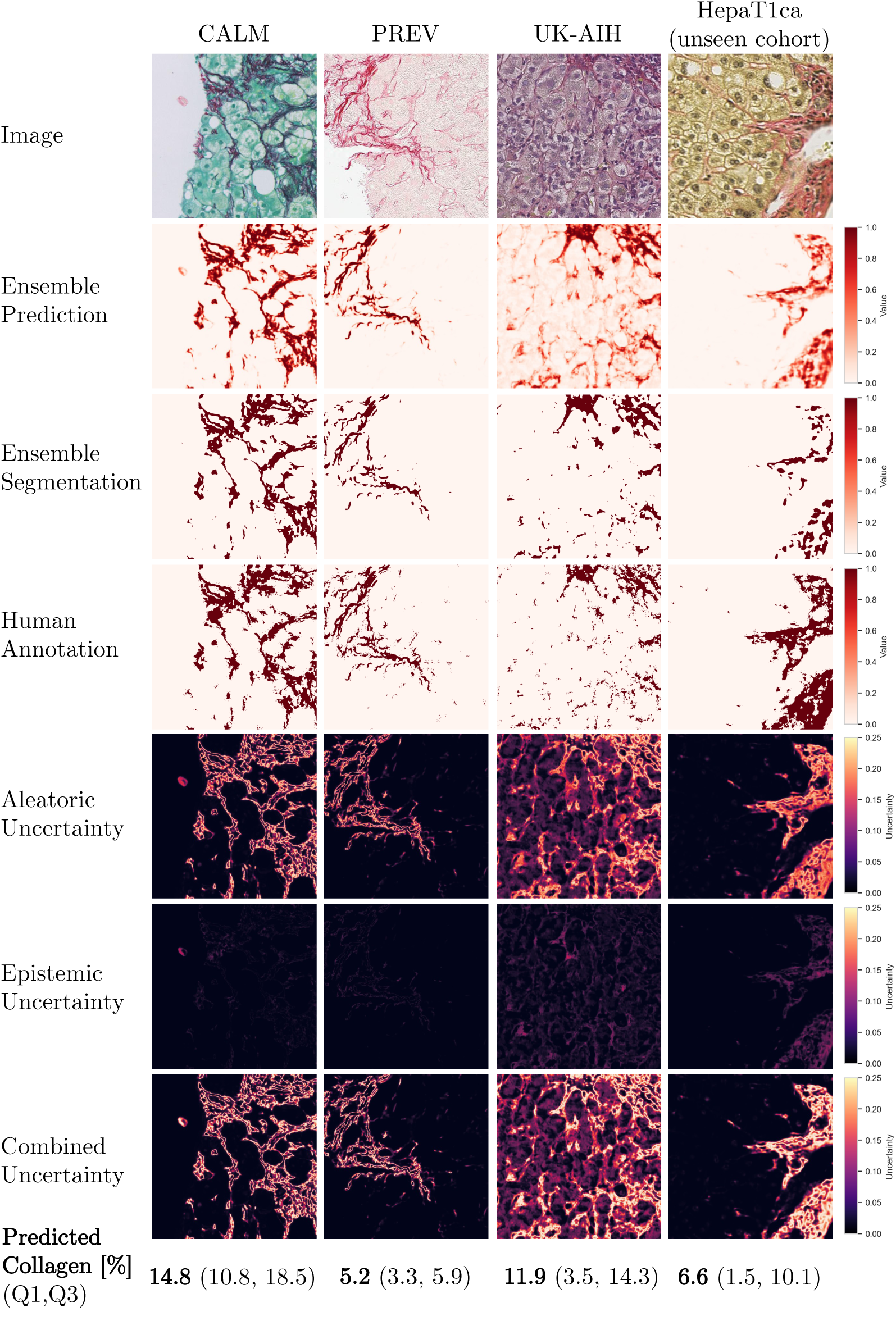
The results of U-Net Mini ensemble model prediction with uncertainty estimations for examples of unseen tiles from each of the cohorts. Tiles with low contrast of stained collagen and unseen colours are characterised by high ensemble model uncertainty.

**Figure 11:**
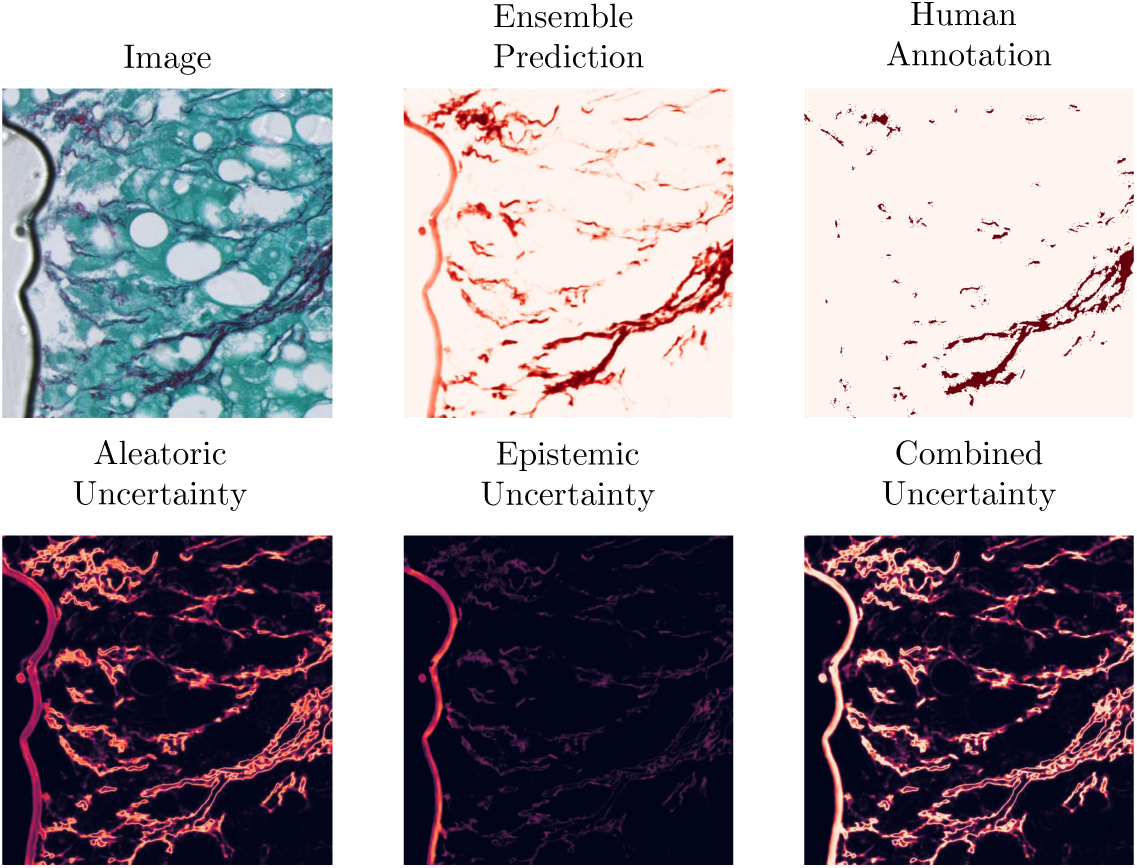
An example image prediction tile with high epistemic uncertainty. The air bubble visible in the original tile is highlighted in the epistemic uncertainty map, which visualises out of distribution data.

Fig. 10 shows the results of model predictions and uncertainty maps for the U-Net Mini ensemble trained on the pooled cohort. It can be seen that almost all of the aleatoric uncertainty is concentrated on the boundaries of collagen fibres. This is expected, as boundary pixels are commonly affected by partial volume effects. Because of this phenomenon, the aleatoric uncertainty is directly tied to the length of the collagen outline in the tile. We observe that tiles with little to no collagen are characterised by uncertainty values near to 0 and tiles with much collagen have high inherent collagen area uncertainty. Uncertainty is computed for all inputs, including slides of suboptimal quality, as part of assessing model robustness under realistic multi-centre conditions. Rather than serving as a rejection criterion, the uncertainty maps indicate which regions within a slide may yield unreliable measurements, complementing rather than replacing conventional quality control.

Table 6 presents the mean ensemble uncertainty scores obtained from model predictions on the validation set. It can be seen that the values of epistemic uncertainty are very low compared to the values of aleatoric uncertainty for all of the image ensembles. We conclude that the uncertainty of the collagen area measurement primarily depends on the characteristics of a given image. The variability of the measured collagen percentage due to differences in predictions by individual segmentation models within each ensemble is relatively very small.

**Table 6:** Ensemble uncertainty scores on the validation set for each of the trained model types. (M = 10 ensemble average ± standard deviation).

| Model ensemble | U Aleatoric | U Epistemic | U Combined |
| --- | --- | --- | --- |
| U-Net Tiny | $0.020 \pm 0.025$ | $0.002 \pm 0.003$ | $0.022 \pm 0.027$ |
| U-Net Mini | $0.017 \pm 0.022$ | $0.002 \pm 0.003$ | $0.019 \pm 0.025$ |
| Att U-Net | $0.024 \pm 0.036$ | $0.003 \pm 0.006$ | $0.027 \pm 0.041$ |

Figure 11 shows ensemble model prediction and uncertainties for a tile with a common imaging artefact, an air bubble. The bubble can be clearly seen in the epistemic uncertainty map, meaning that some of the models in the ensemble have classified its pixels as regions of collagen, and some did not. The visualisation of epistemic uncertainty can provide the pathologist with a map of regions in the slide which were not represented in the model training set, and therefore are likely to contain prediction errors. These examples illustrate the practical value of uncertainty quantification: regions with high epistemic uncertainty frequently correspond to visible artefacts or staining inconsistencies, indicating a higher likelihood of segmentation errors. While a quantitative correlation between uncertainty magnitude and segmentation accuracy would require ground-truth fibrosis scoring across all cohorts, the qualitative consistency observed in Figures 10 and 11 demonstrates that uncertainty maps effectively highlight potentially unreliable regions. In this way, uncertainty estimation not only improves interpretability but also provides a mechanism to flag regions requiring closer manual inspection.

### 4.2. Interpreting uncertainty

The results of uncertainty scores computed for each of the cohorts are provided in Table 7. The PREV cohort has the lowest aleatoric uncertainty values of the cohorts studied. The aleatoric uncertainty is highly correlated to the area of collagen in an image (see Fig. A.13) and therefore the lower uncertainty in the PREV cohort can be partly explained by the lower content of collagen in this cohort overall (see Table 3).

**Table 7:** Ensemble prediction uncertainty scores for validation samples from each of the cohorts, U-Net Mini. (M = 10 ensemble average ± standard deviation).

| Training Set | CALM | PREV | UK-AIH | HepaT1ca |
| --- | --- | --- | --- | --- |
| **Individual Cohorts** |  |  |  |  |
| U Aleatoric | $0.016 \pm 0.016$ | $0.006 \pm 0.008$ | $0.024 \pm 0.020$ | - |
| U Epistemic | $0.002 \pm 0.003$ | $0.001 \pm 0.003$ | $0.003 \pm 0.005$ | - |
| **Pooled Cohort** |  |  |  |  |
| U Aleatoric | $0.026 \pm 0.028$ | $0.008 \pm 0.011$ | $0.044 \pm 0.033$ | $0.018 \pm 0.022$ |
| U Epistemic | $0.002 \pm 0.002$ | $0.001 \pm 0.001$ | $0.001 \pm 0.005$ | $0.002 \pm 0.004$ |

Interestingly, after cohort pooling the mean aleatoric uncertainty scores increase for all the models, especially in the cohorts where more than one stain colour was used (i.e. CALM and UK-AIH). This indicates that the segmentation models introduced to different variants of collagen staining have lower confidence at the collagen boundaries where more than one stain is present. Such interpretation would correspond to the intuition that placing additional stains on the tissue specimen introduces additional sources of variability in the data. The highest aleatoric uncertainty values are reported for the UK-AIH cohort in which the purple haematoxylin staining was used in the PSR slides. Collagen tiles in this cohort have overall a lower visual contrast than those from other cohorts.

It can be seen that the correlation between the average prediction *P* and the segmented collagen *B* is equal to 1, meaning that these measures can be considered equivalent to each other (Fig. A.13). This means that it may be acceptable to present pathologists with the collagen prediction maps rather than the binarised segmentation maps. The prediction maps have a closer resemblance to the original input image as they capture the colour intensity gradients characteristic of collagen fibres of different thicknesses. Collagen prediction maps are also visually more similar to the images acquired using SHG/TPEF microscopy, where similarly, an inferred collagen density map rather than a binary segmentation map is obtained [49]. Such continuous collagen prediction maps could be interpreted as a kind of “ideal” stain deconvolution, where only pixels containing any collagen can have positive values, and regions of tissue which have no collagen (i.e. cell cytoplasm, cell nuclei) have a collagen probability of 0.

From a practical standpoint, the acceptable level of uncertainty will depend on the intended use of the segmentation output. Based on the ensemble statistics in Table 7, epistemic uncertainty values for the U-Net Mini trained on the pooled cohort were typically around 0.002. Predictions with markedly higher epistemic uncertainty may indicate out-of-distribution image regions or unreliable model behaviour and should be prioritised for review. Aleatoric uncertainty, by contrast, reflects intrinsic image ambiguity (for example, collagen fibre boundaries) and is therefore less suitable for defining reliability thresholds. In practice, uncertainty maps can serve as an intuitive visual cue to guide users toward regions that may warrant closer inspection.

## 5. Discussion

The problem of performing quantitative histological analysis on differently stained slides is well recognised. Early stain normalisation relied on colour deconvolution [6, 7, 50], but its assumptions of white-light illumination and stable stain vectors often fail in whole-slide imaging [9]. Later, colour-transfer and generative approaches, including histogram matching [10], sparse autoencoders [11], and CycleGAN-based models [12–16], achieved visually realistic transformations, yet their biological fidelity remains uncertain.

While domain adaptation strategies, including stain normalisation and colour transfer, have been proposed to improve cross-site generalisability, their impact on prediction and segmentation accuracy remains limited [51, 52]. In practice, deep ensembles trained on diverse data may offer greater robustness, particularly when supported by uncertainty quantification. Nonetheless, exploring domain adaptation remains a potential direction for future work in settings with limited training diversity or highly divergent staining.

Automated quality-control tools such as HistoQC are highly effective for detecting slide-level artefacts (e.g., tissue folds, pen marks, out-of-focus regions) and can also report basic colour and intensity statistics [17]. However, these tools are primarily designed for identifying outliers or artefacts rather than systematically quantifying inter-cohort staining variability. Our colour-space analysis therefore provides a complementary perspective, enabling cohort-level assessment of staining and acquisition differences. Future studies may benefit from combining colour-space characterisation with automated QC metrics to provide both global and local assessments of data quality.

Beyond quality control, uncertainty estimation offers an orthogonal way to assess the reliability of automated predictions. Uncertainty estimates can inform data-curation workflows by directing human annotation toward the most ambiguous image regions, supporting active-learning paradigms that improve efficiency without compromising quality. In this context, uncertainty estimation should not be viewed as a replacement for quality control or data curation, but as a complementary mechanism for assessing the trustworthiness of model predictions on heterogeneous, real-world data. By quantifying prediction confidence directly from the model output, uncertainty maps provide a means to contextualise results even when input quality varies across centres.

While the U-Net architecture is long established in biomedical image segmentation, it remains a robust reference model valued for its stability, interpretability, and extensive validation across histopathology tasks. Our aim in this study was not to benchmark architectures, but to evaluate the estimation and interpretation of predictive uncertainty. Using the U-Net provided a controlled and transparent framework for ensemble-based uncertainty analysis across diverse staining conditions. We acknowledge that newer architectures, including transformer-based and foundation-model approaches, may further improve cross-site generalisation [53, 54]. However, our results show that even this lightweight and classical model achieves strong multi-centre performance (Dice 0.83–0.90) with modest computational cost, suggesting that uncertainty estimation itself—rather than architectural novelty—offers substantial benefits for model interpretability and reliability.

From a computational standpoint, ensemble-based uncertainty estimation inevitably increases training and inference time compared with a single model. However, the overhead is moderate: inference of a 512 × 512 px tile required approximately 2.5 seconds on a Quadro RTX 6000 GPU (A.8). Given the interpretability and calibration advantages of ensemble methods, this cost is acceptable for research and batch-processing workflows, particularly in settings where throughput is less critical than reliability.

The extreme staining variability observed across cohorts—particularly in the number and type of counterstains—highlights the challenge of developing a unified segmentation approach under real-world conditions. Rather than excluding these divergent slides, we deliberately retained them to assess how ensemble-based uncertainty estimation performs across heterogeneous inputs. Our findings indicate that, despite substantial colour variation, the model maintained acceptable cross-cohort performance, and uncertainty maps successfully identified regions where predictions were less reliable. This suggests that uncertainty estimation can complement stain harmonisation efforts by providing a mechanism to interpret and manage variability, while future work should aim to integrate these strategies through domain adaptation or foundation-model frameworks.

Although non-invasive imaging methods continue to advance [55], histology remains essential for definitive fibrosis assessment. By improving the reliability and interpretability of slide-based quantification, our approach supports this diagnostic foundation and facilitates comparison with emerging non-invasive metrics.

## 6. Conclusions

This study demonstrates that ensemble-based uncertainty estimation strengthens quality control and supports more reliable deployment of automated collagen quantification in multi-centre liver biopsy cohorts. Across four independent PSR-stained cohorts, we observed pronounced staining heterogeneity. Within this setting, the ensemble U-Net achieved strong segmentation performance in the training cohorts (Dice 0.83–0.90) and generated informative uncertainty maps. These maps captured both image-intrinsic ambiguity and model-related uncertainty, allowing direct visualisation of prediction confidence at the pixel level. Segmentation performance decreased in the unseen HepaT1ca cohort, reflecting substantial staining divergence. The uncertainty maps nevertheless remained spatially informative and preserved interpretability of model behaviour under domain shift.

The combined use of ensemble prediction, uncertainty mapping, and inter-cohort colour-distribution analysis enabled transparent evaluation of model behaviour across centres. Explicit quantification of prediction confidence extends validation beyond aggregate performance metrics and supports informed interpretation of automated collagen proportionate-area measurements. In practice, epistemic uncertainty provides a mechanism for identifying unreliable predictions and prioritising them for expert review, reinforcing existing quality-control procedures.

Future multi-centre studies will benefit from incorporating structured characterisation of stain differences and systematic analysis of uncertainty-associated failure modes alongside conventional performance metrics. Integration of such evaluation strategies with continued stain harmonisation efforts and foundation-model representations trained on diverse histopathology data will further strengthen cross-centre reliability. Collectively, these approaches advance the development of more transparent, trustworthy, and reproducible computational pathology workflows.

## Data Availability

Data produced in the present study are available upon reasonable request to the authors.

## Acknowledgements

The computational aspects of this research were supported by the Wellcome Trust Core Award Grant Number 203141/Z/16/Z and the NIHR Oxford BRC. The views expressed are those of the author(s) and not necessarily those of the NHS, the NIHR or the Department of Health.

We are indebted to all the members of the clinical and research teams in centres who have recruited to UK-AIH and are key to the ongoing success of the study. The principal investigators and recruiting centres are: Dr Shirley English (Aberdeen Royal Infirmary), Dr Graeme Alexander/Dr George Mells (Addenbrooke’s Hospital), Dr Debabrata Majumdar (St Peters Hospital), Dr Vinay Sathyanarayana (Barnsley Hospital), Professor John Ramage (Basingstoke North Hampshire Hospital), Dr Christopher Shorrock (Blackpool Victoria Hospital), Dr James Maggs (Buckinghamshire Hospital), Dr David Elphick (Chesterfield Royal Hospital), Dr Chris Macdonald (Cumberland Infirmary), Professor Matthew Cramp (Derriford Hospital), Dr Joanne Sayer (Doncaster Royal Infirmary), Dr James Jupp (Dorset County Hospital), Dr Jessica Dyson (Freeman Hospital), Dr Coral Hollywood (Gloucestershire Royal Hospital), Dr Alexandra Daley (Heartlands Hospital), Dr Lynsey Corless (Hull Royal Infirmary), Dr Darren Craig (James Cook University Hospital), Dr Emma Culver (John Radcliffe Hospital, Oxford), Professor Michael Heneghan (King’s College Hospital), Dr Sharat Misra (King’s Mill Hospital), Dr Chris Corbett (New Cross Hospital), Professor John Dillon (Ninewells Hospital), Dr Simon Rushbrook (Norfolk and Norwich University Hospital), Dr Thomas Lee (North Tyneside General Hospital), Dr Nicholas M Sharaer (Poole Hospital), Dr Kara Rye (Princess Royal Hospital), Dr Andrew Fowell (Queen Alexandra Hospital, Portsmouth), Dr Andrea Broad/Dr Dina Mansour (Queen Elizabeth Hospital, Gateshead), Dr Andy Douds (Queen Elizabeth Hospital, King’s Lynn), Dr Stephen Ryder (Queen’s Medical Centre), Dr Richard Keld (Royal Albert Edward Infirmary), Dr Earl Williams (Royal Bournemouth Hospital), Dr William Stableforth (Royal Cornwall Hospital), Dr Andrew Austin (Royal Derby Hospital), Professor Dermot Gleeson (Royal Hallamshire Hospital), Dr Kenneth Simpson (Royal Infirmary of Edinburgh), Dr Imran Patanwala (Royal Liverpool University Hospital), Dr Alison Brind (Royal Stoke University Hospital), Dr Shanika de Silva (Russells Hall Hospital), Dr Aqueel Jamil (Salisbury District Hospital), Dr Saket Singhal (Sandwell General Hospital), Dr Chin Lye Ch’ng (Singleton Hospital), Dr Joanne Topping (South Tyneside District Hospital), Dr Mark Wright (Southampton General Hospital), Dr Talal Valliani (Southmead Hospital), Dr Rebecca Jones (St. James’s University Hospital, Leeds), Dr Harriet Mitchison (Sunderland Royal Hospital), Dr Douglas Thorburn (The Royal Free Hospital), Professor Aftab Ala (The Royal Surrey County Hospital), Dr Ye Htun Oo (University Hospital Birmingham NHS Foundation Trust), Dr Sushma Sakena/Dr Francisco Porras-Perez (University Hospital of North Durham), Prof Jane Metcalf/Dr Stephen Mitchell (University Hospital of North Tees), Dr Esther Unitt/Dr Victoria Gordon (University Hospitals Coventry Warwick), and Dr Jeremy Shearman (Warwick Hospital, South Warwickshire NHS Foundation Trust).

JKD is supported by the NIHR Newcastle Biomedical Research Centre. The UK-AIH study is funded by LiverNorth (a national patient charity) and previously by the NIHR Rare Diseases Translational Research Collaboration. The views expressed are those of the author(s) and not necessarily those of the NHS, the NIHR or the Department of Health.

## 8. Code Availability

The source code for the slide colour analysis tool is available under: github.com/mkatw/slide colour palette.

## Appendix A. Appendix

### Appendix A.1. Processing time

Table A.8 shows a comparison of the training and inference times for each of the tested models. The times are reported for the processing of the training set (training over 30 epochs) and of the validation set, respectively. It can be seen that tile processing time is strongly related to the size of the network used, with U-Net Tiny outperforming other models in our study. Processing time is an important factor to consider when choosing deep learning models for use in clinical practice.

**Table A.8:** Training and inference time comparison for the trained models. (M = 10 ensemble average ± standard deviation).

| Model | Training [s] | Inference [s] |
| --- | --- | --- |
| U-Net Tiny | $417.7 \pm 16.0$ | $1.3 \pm 0.2$ |
| U-Net Mini | $1358.8 \pm 24.5$ | $2.5 \pm 0.6$ |
| Att U-Net | $1590.4 \pm 37.0$ | $3.3 \pm 0.8$ |

### Appendix A.2. Correlation statistics

Figures A.12 and A.13 show the correlation statistics for computed collagen areas (the average prediction *P* and the segmented collagen area *B*), as well as uncertainties for the models trained on the individual cohorts and the model trained on the pooled cohort, respectively.

**Figure A.12:**
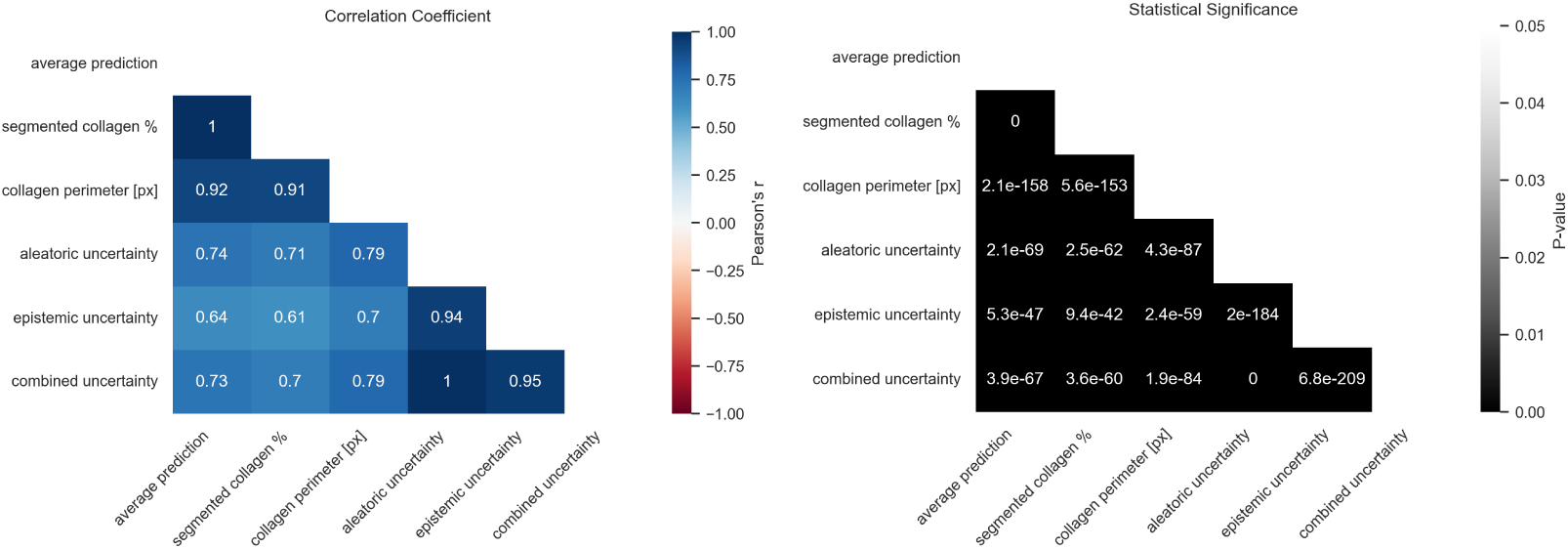
Correlation coefficients and corresponding statistical significance values for the prediction parameters of the model trained on the individual cohorts. The values are reported for predictions obtained using the U-Net Mini ensemble.

**Figure A.13:**
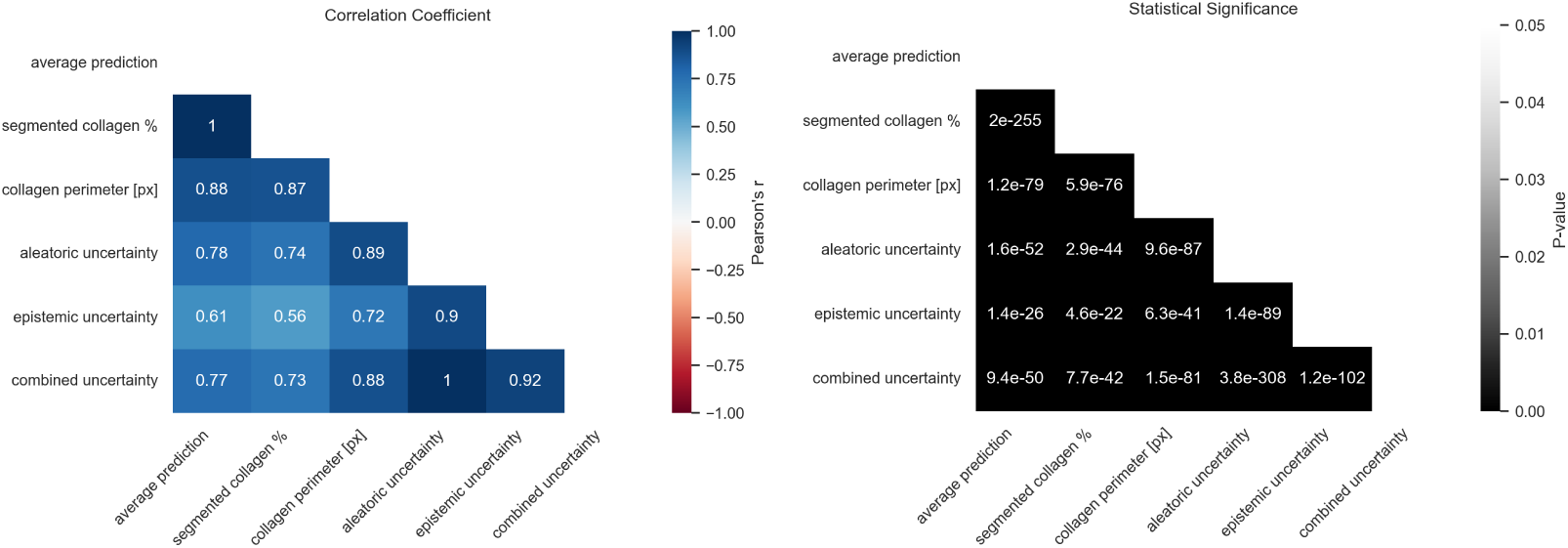
Correlation coefficients and corresponding statistical significance values for the prediction parameters of the model trained on the pooled cohort. The values are reported for predictions obtained using the U-Net Mini ensemble.

## Footnotes

1 https://github.com/stefano-malacrino/wsi-reader

2 https://github.com/cgohlke/tifffile

3 https://github.com/yingkaisha/keras-unet-collection

## References

[1] Roberta Forlano et al. “NAFLD: Time to apply quantitation in liver biopsies as endpoints in clinical trials”. In: Journal of Hepatology 74.1 (2021), pp. 241–242.

2. Elena Buzzetti, et al. “Collagen proportionate area is an independent predictor of long-term outcome in patients with non-alcoholic fatty liver disease”. In: Alimentary Pharmacology and Therapeutics 49.9 (2019), pp. 1214–1222.

3. Mads Israelsen, et al. “Collagen proportionate area predicts clinical outcomes in patients with alcohol-related liver disease”. In: Alimentary Pharmacology and Therapeutics 52.11-12 (2020), pp. 1728–1739.

4. Amanda Cheung, et al. “Defining Improvement in Nonalcoholic Steatohepatitis for Treatment Trial Endpoints: Recommendations From the Liver Forum”. In: Hepatology 70.5 (2019), pp. 1841–1855.

5. Alexandros Arjmand, et al. “Quantification of liver fibrosis-a comparative study”. In: Applied Sciences (Switzerland) 10.2 (2020), pp. 1–34.

6. A. C. Ruifrok and D. A. Johnston. “Quantification of histochemical staining by color deconvolution”. In: Analytical and Quantitative Cytology and Histology 23.4 (2001), pp. 291–299.

[7] Marc Macenko et al. “A method for normalizing histology slides for quantitative analysis”. In: Proceedings 2009 IEEE International Symposium on Biomedical Imaging: From Nano to Macro, ISBI 2009 (2009), pp. 1107–1110.

[8] Abhishek Vahadane et al. “Structure-preserved color normalization for histological images”. In: Proceedings International Symposium on Biomedical Imaging 2015-July (2015), pp. 1012–1015.

9. Peter Haub and Tobias Meckel. “A model based survey of colour deconvolution in diagnostic brightfield microscopy: Error estimation and spectral consideration”. In: Scientific Reports 5.February (2015), pp. 1– 15.

10. Erik Reinhard, et al. “Color transfer between images”. In: IEEE Computer Graphics and Applications 21.5 (2001), pp. 34–41.

11. Andrew Janowczyk, Ajay Basavanhally, and Anant Madabhushi. “Stain Normalization using Sparse AutoEncoders (StaNoSA): Application to digital pathology”. In: Computerized Medical Imaging and Graphics 57 (2017), pp. 50–61.

12. Jun-yan Zhu, et al. “Unpaired Image-to-Image Translation using Cycle-Consistent Adversarial Networks”. In: (2017). arXiv: 1703.10593.

[13] M. Tarek Shaban et al. “Staingan: Stain style transfer for digital histological images”. In: Proceedings International Symposium on Biomedical Imaging 2019-April (2019), pp. 953–956.

14. Yair Rivenson, et al. “PhaseStain: the digital staining of label-free quantitative phase microscopy images using deep learning”. In: Light: Science and Applications 8.1 (2019). arXiv: 1807.07701.

[15] Kevin de Haan, et al. “Deep learning-based transformation of H&E stained tissues into special stains”. In: Nature Communications 12.1 (2021), pp. 1–13.

[16] Nicolas Wagner et al. Federated Stain Normalization for Computational Pathology. Vol. 1. Springer Nature Switzerland, 2022, pp. 14–23.

[17] Andrew Janowczyk et al. “HistoQC: An Open-Source Quality Control Tool for Digital Pathology Slides”. In: JCO Clinical Cancer Informatics (3 Dec. 2019), pp. 1–7. url: https://ascopubs.org/doi/10.1200/ CCI.18.00157.

[18] Yoshio Sumida, Atsushi Nakajima, and Yoshito Itoh. Limitations of liver biopsy and non-invasive diagnostic tests for the diagnosis of nonalcoholic fatty liver disease/nonalcoholic steatohepatitis. 2014.

[19] Qiang Li et al. “Virtual liver needle biopsy from reconstructed threedimensional histopathological images: Quantification of sampling error”. In: Computers in Biology and Medicine 147 (Aug. 2022).

[20] Timothy J. Kendall, et al. “Hepatic elastin content is predictive of adverse outcome in advanced fibrotic liver disease”. In: Histopathology 73.1 (2018), pp. 90–100.

[21] Holde Puchtler and Faye Sweat. “Histochemical specifity of staining methods for connective tissue fibers: Resorcin-fuchsin and van Gieson’s picro-fuchsin”. In: Histochemie 4 (1964), pp. 24–34.

[22] R. W. Dapson, et al. “Certification procedures for sirius red F3B (CI 35780, Direct red 80)”. In: Biotechnic and Histochemistry 86.3 (2011), pp. 133–139.

[23] Stéphane Bancelin, et al. “Determination of collagen fibril size via absolute measurements of second-harmonic generation signals”. In: Nature Communications 5 (2014), pp. 1–8.

[24] Seyed Mohammad Siadat et al. “Measuring collagen fibril diameter with differential interference contrast microscopy”. In: Journal of Structural Biology 213.1 (2021), p. 107697.

[25] Benoit B Mandelbrot. *The fractal geometry of nature*. eng. [Rev. ed.] New York: W.H. Freeman, 1983.

[26] Nicola Dioguardi et al. “Liver fibrosis and tissue architectural change measurement using fractal-rectified metrics and Hurst’s exponent”. In: World Journal of Gastroenterology 12.14 (2006), pp. 2187–2194.

[27] Ender Gunes Yegin, Korkut Yegin, and Osman Cavit Ozdogan. “Digital image analysis in liver fibrosis: basic requirements and clinical implementation”. In: Biotechnology & Biotechnological Equipment 30.4 (2016), pp. 653–660.

28. Cyrill Matenaers, et al. “Practicable methods for histological section thickness measurement in quantitative stereological analyses”. In: PLoS ONE 13.2 (2018), pp. 1–21.

[29] Stuart Astbury et al. “Reliable computational quantification of liver fibrosis is compromised by inherent staining variation”. In: Journal of Pathology: Clinical Research 7.5 (2021), pp. 471–481.

[30] Natasha Mcdonald, et al. “Multiparametric magnetic resonance imaging for quantitation of liver disease: a two-centre crosssectional observational study”. In: Scientific Reports February (2018), pp. 1–10.

31. Damian J. Mole, et al. “Study protocol: HepaT1ca An observational clinical cohort study to quantify liver health in surgical candidates for liver malignancies”. In: BMC Cancer 18.1 (2018), pp. 1–7.

[32] Stephen A. Harrison et al. “Prospective evaluation of the prevalence of non-alcoholic fatty liver disease and steatohepatitis in a large middleaged US cohort”. In: Journal of Hepatology 75.2 (2021), pp. 284–291.

33. Jessica K. Dyson, et al. “Inequity of care provision and outcome disparity in autoimmune hepatitis in the United Kingdom”. In: Alimentary Pharmacology and Therapeutics 48.9 (2018), pp. 951–960.

[34] Adam Goode et al. “OpenSlide: A vendor-neutral software foundation for digital pathology”. In: Journal of Pathology Informatics 4.1 (2013), p. 27.

35. Peter Bankhead, et al. “QuPath: Open source software for digital pathology image analysis”. In: Scientific Reports 7.1 (2017), p. 16878.

[36] Paul Heckbert. “Color Image Quantization for Frame Buffer Display”. In: Computer Graphics 16.3 (1982), pp. 157–166.

[37] Nobuyuki Otsu. “A Threshold Selection Method from Gray-Level Histograms”. In: IEEE Transactions on Systems, Man, and Cybernetics C.1 (1979), pp. 62–66.

[38] Stéfan Van Der Walt et al. “Scikit-image: Image processing in python”. In: *PeerJ* 2014.1 (2014), pp. 1–18.

[39] Olaf Ronneberger, Philipp Fischer, and Thomas Brox. U-Net: Convolutional Networks for Biomedical Image Segmentation. Ed. by Nassir Navab et al. Vol. 9351. Lecture Notes in Computer Science. Cham: Springer International Publishing, 2015, pp. 234–241.

40. Ozan Oktay, et al. “Attention U-Net: Learning Where to Look for the Pancreas”. In: Midl (2018). arXiv: 1804.03999.

[41] Yarin Gal and Zoubin Ghahramani. “Dropout as a Bayesian Approximation: Representing Model Uncertainty in Deep Learning”. In: Proceedings of The 33rd International Conference on Machine Learning. Ed. by Maria Florina Balcan and Kilian Q. Weinberger. Vol. 48. Proceedings of Machine Learning Research. New York, New York, USA: PMLR, 2016, pp. 1050–1059.

[42] Alex Kendall and Yarin Gal. “What Uncertainties Do We Need in Bayesian Deep Learning for Computer Vision?” In: Advances in Neural Information Processing Systems. Ed. by I. Guyon et al. Vol. 30. Curran Associates, Inc., 2017.

[43] Radford M Neal. Bayesian Learning for Neural Networks. 1996.

[44] Alex Graves. “Practical Variational Inference for Neural Networks”. In: Advances in Neural Information Processing Systems. Ed. by J. Shawe-Taylor et al. Vol. 24. Curran Associates, Inc., 2011.

[45] Balaji Lakshminarayanan, Alexander Pritzel, and Charles Blundell. “Simple and Scalable Predictive Uncertainty Estimation Using Deep Ensembles”. In: Proceedings of the 31st International Conference on Neural Information Processing Systems. NIPS’17. Long Beach, California, USA: Curran Associates Inc., 2017, pp. 6405–6416.

[46] Armen Der Kiureghian and Ove Ditlevsen. “Aleatory or epistemic? Does it matter?” In: Structural Safety 31.2 (2009). Risk Acceptance and Risk Communication, pp. 105–112.

[47] Yongchan Kwon, et al. “Uncertainty quantification using Bayesian neural networks in classification: Application to biomedical image segmentation”. In: Computational Statistics and Data Analysis 142 (2020), p. 106816.

48. Stanislav Fort, Huiyi Hu, and Balaji Lakshminarayanan. “Deep Ensembles: A Loss Landscape Perspective”. In: (2019), pp. 1–15. arXiv: 1912.02757.

[49] Shuoyu Xu et al. “QFibrosis: A fully-quantitative innovative method incorporating histological features to facilitate accurate fibrosis scoring in animal model and chronic hepatitis B patients”. In: Journal of Hepatology 61.2 (2014), pp. 260–269.

50. Adnan Mujahid Khan, et al. “A nonlinear mapping approach to stain normalization in digital histopathology images using image-specific color deconvolution”. In: IEEE Transactions on Biomedical Engineering 61.6 (2014), pp. 1729–1738.

[51] Wingates Voon et al. “Evaluating the effectiveness of stain normalization techniques in automated grading of invasive ductal carcinoma histopathological images”. In: Scientific Reports 13 (1 Dec. 2023).

[52] Amirreza Mahbod et al. “Improving generalization capability of deep learning-based nuclei instance segmentation by non-deterministic train time and deterministic test time stain normalization”. In: Computational and Structural Biotechnology Journal 23 (Dec. 2024), pp. 669– 678.

[53] Jieneng Chen et al. “TransUNet: Rethinking the U-Net architecture design for medical image segmentation through the lens of transformers”. In: Medical Image Analysis 97 (Oct. 2024), p. 103280.

[54] Richard J. Chen et al. “Towards a general-purpose foundation model for computational pathology”. In: Nature Medicine 30 (3 Mar. 2024), pp. 850–862.

55. Richard K. Sterling, et al. “AASLD Practice Guideline on imaging-based noninvasive liver disease assessment of hepatic fibrosis and steatosis”. In: Hepatology (2024).

